# Mass incarceration as a driver of the tuberculosis epidemic in Latin America and projected impacts of policy alternatives: A mathematical modeling study

**DOI:** 10.1101/2024.04.23.24306238

**Authors:** Yiran E Liu, Yasmine Mabene, Sergio Camelo, Zulma Vanessa Rueda, Daniele Maria Pelissari, Fernanda Dockhorn Costa Johansen, Moises A Huaman, Tatiana Avalos-Cruz, Valentina A Alarcón, Lawrence M Ladutke, Marcelo Bergman, Ted Cohen, Jeremy D Goldhaber-Fiebert, Julio Croda, Jason R Andrews

## Abstract

**Background:** Tuberculosis incidence is increasing in Latin America, where the incarcerated population has nearly quadrupled since 1990. The full impact of incarceration on the tuberculosis epidemic, accounting for effects beyond prisons, has never been quantified.

**Methods:** We calibrated dynamic compartmental transmission models to historical and contemporary data from Argentina, Brazil, Colombia, El Salvador, Mexico, and Peru, which comprise approximately 80% of the region’s incarcerated population and tuberculosis burden. Using historical counterfactual scenarios, we estimated the transmission population attributable fraction (tPAF) for incarceration and the excess population-level burden attributable to increasing incarceration prevalence since 1990. We additionally projected the impact of alternative incarceration policies on future population tuberculosis incidence.

**Findings:** Population tuberculosis incidence in 2019 was 29.4% (95% UI, 23.9-36.8) higher than expected without the rise in incarceration since 1990, corresponding to 34,393 (95% UI, 28,295-42,579) excess incident cases across countries. The incarceration tPAF in 2019 was 27.2% (95% UI, 20.9-35.8), exceeding estimates for other risk factors like HIV, alcohol use disorder, and undernutrition. Compared to a scenario where incarceration rates remain stable at current levels, a gradual 50% reduction in prison admissions and duration of incarceration by 2034 would reduce population tuberculosis incidence by over 10% in all countries except Mexico.

**Interpretation:** The historical rise in incarceration in Latin America has resulted in a large excess tuberculosis burden that has been under-recognized to-date. International health agencies, ministries of justice, and national tuberculosis programs should collaborate to address this health crisis with comprehensive strategies, including decarceration.

**Funding:** National Institutes of Health

**Research in context:** *Evidence before this study:* We searched PubMed for studies on tuberculosis in prisons in Latin America, using the search terms (“tuberculosis”) AND (“prisons” OR “incarceration”) AND (“Latin America” OR “Argentina” OR “Brazil” OR “Colombia” OR “El Salvador” OR “Mexico” OR “Peru”), published in any language. Previous studies have identified a high risk of tuberculosis in prisons in Latin America, finding that notifications in prisons are increasing and account for a growing proportion of all cases in the region. Other national or sub-national studies have found elevated tuberculosis risk among formerly incarcerated individuals and transmission chains spanning prisons and communities. However, the full contribution of incarceration to the broader tuberculosis epidemic in Latin America—accounting for historical incarceration trends, under-detection in prisons, and “spillover” effects into communities—has never been quantified. Furthermore, previous studies have evaluated biomedical interventions in prisons; the regional impact of alternative incarceration policies on future population tuberculosis incidence is unknown.

*Added value of this study:* Here we quantify the full contribution of incarceration to the tuberculosis epidemic in Latin America. Our model captures the dynamic nature of incarceration, incorporating historical and contemporary data sources to account for varying prison turnover rates and mechanisms underlying historical incarceration growth. By modeling the population with incarceration history, we estimate the true size of the ever-exposed population, which across the six countries is over 11 times the size of the population within prison at any one time. We identify the settings where excess cases occur and compare our results to crude estimates based on notifications in prisons. We show, across six countries with diverse carceral contexts and tuberculosis epidemiology, that incarceration is a leading driver on par with other major tuberculosis risk factors, a role that has been under-recognized to date. Finally, we demonstrate the potential impact of alternative incarceration policies in reducing future tuberculosis burden in carceral settings and the general population.

*Implications of all the available evidence:* To date the true impact of incarceration on the tuberculosis epidemic across the region has been underestimated due to a narrow focus on disease occurring during incarceration. In light of the substantial excess tuberculosis burden attributable to incarceration, interventions targeting incarceration can have outsized effects on the broader tuberculosis epidemic in Latin America— much greater than previously appreciated. These interventions should include not only strategies to reduce tuberculosis risk among currently and formerly incarcerated individuals, but also efforts to end mass incarceration.

## INTRODUCTION

Globally, 10.6 million people developed tuberculosis in 2022^1^. While global tuberculosis incidence has decreased by 8.7% since 2015, in Latin America, tuberculosis incidence increased by 19% over the same period, highlighting the urgent need to address key tuberculosis drivers in the region.

In Latin America, the incarcerated population has nearly quadrupled over the last thirty years, the most rapid growth of any region in the world^2^. Persons deprived of liberty (PDL), who may already face elevated risk of tuberculosis prior to incarceration, are further exposed to prison conditions that foster transmission and disease progression, including overcrowding, poor ventilation, malnourishment, and limited access to health care^3^. Together these factors contribute to tuberculosis rates that, in South America, are 26 times higher among PDL than in the general population^4^.

Recognizing the crisis of tuberculosis in prisons, the Pan-American Health Organization (PAHO) began requesting data from member states on case notifications occurring among PDL. Between 2014 and 2019, the percent of all notified tuberculosis cases in the region occurring among PDL increased from 6.6% to 9.4%^3,5^. While alarmingly high, this figure underestimates the tuberculosis burden attributable to incarceration, for several reasons. First, the case detection ratio is lower in prisons than in the general population^5^. Second, individuals who acquire infection in prison often do not progress to tuberculosis disease until after release. Indeed, previous studies showed that formerly incarcerated individuals had elevated rates of tuberculosis for up to seven years following release from prison^6,7^. As notifications databases do not record information on incarceration history, these cases are not currently attributed to incarceration^8^. Finally, infections acquired in prisons, including among people who work in or visit prisons, can spread in the community. Accordingly, genomic epidemiologic studies have identified tuberculosis transmission chains that span prisons and communities^9–13^. Therefore, existing studies that focus on tuberculosis occurring in prisons overlook the role of incarceration as a population-level tuberculosis driver.

Understanding the full contribution of incarceration to the worsening tuberculosis epidemic in Latin America is critical to inform tuberculosis prevention strategies and resource allocation. Furthermore, the impact of alternative incarceration policies on the tuberculosis epidemic remains unknown, as previous studies have focused on biomedical interventions. In this study, we use mathematical modeling to quantify the population-level burden of tuberculosis attributable to incarceration in six countries: Argentina, Brazil, Colombia, El Salvador, Mexico, and Peru. Specifically, we hypothesize that the rise in incarceration since 1990 has produced a growing excess tuberculosis burden and hindered tuberculosis progress in the region. We additionally simulate alternative incarceration policies and project their impact on future population tuberculosis incidence.

## METHODS

### Setting

We selected countries in Latin America, defined as Mexico, Central America, and South America, based on data availability and to represent regional heterogeneity in incarceration and tuberculosis trends (**Figure 1**). Together, the six included countries represent 82.4% of the region’s incarcerated population, 79.7% of total tuberculosis notifications, and 80.1% of tuberculosis notifications in prisons in 2018.

**Figure 1.**
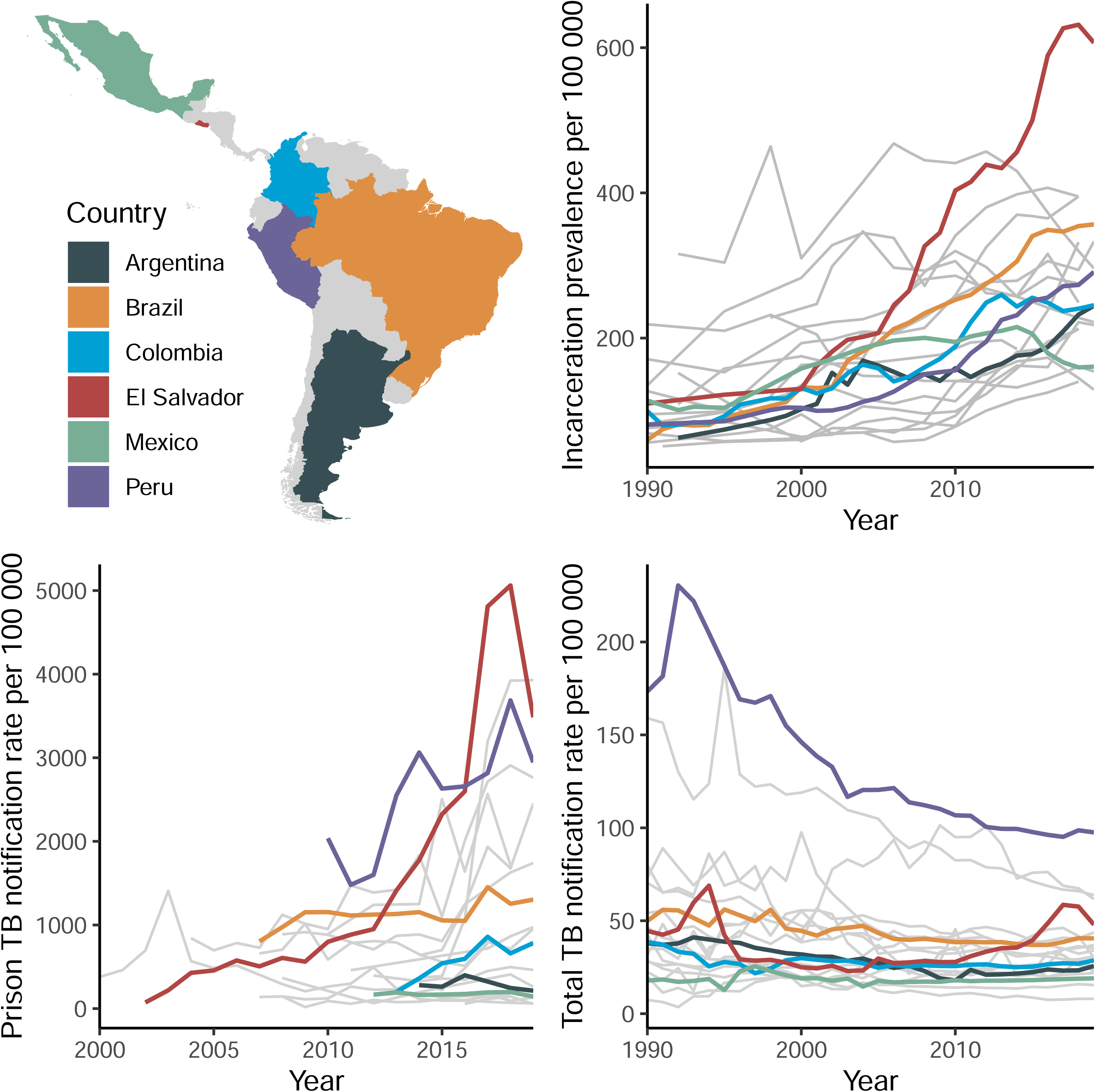
Geographic, demographic, and epidemiologic heterogeneity among included countries. Countries included in the analysis are highlighted in color; remaining countries in Latin America are depicted in grey. Incarceration prevalence refers to the number of people per 100,000 population who are incarcerated at a given point in time. Tuberculosis (TB) notification rates are per 100,000 person-years. Data on prison tuberculosis notifications are only available starting in 2000. Latin America includes Mexico, Central America, and South America.

### Data sources

We collected data on incarceration prevalence, prison entries or releases, and recidivism from each country’s penitentiary department or census agency via published reports and information requests (**Table S1, Appendix**). We also referenced reports and articles published by researchers, international agencies, and journalists. Population estimates and projections were obtained from World Population Prospects. Population-wide tuberculosis notifications and incidence estimates were retrieved from the WHO Global Tuberculosis Report^1^. Notifications and incidence estimates for PDL were sourced from PAHO and a recent study^5^ (**Table S2**).

### Model development and calibration

We developed a deterministic, meta-population compartmental model to simulate incarceration and tuberculosis transmission (**Figure S1**). The model includes a simple representation of tuberculosis natural history across five compartments: susceptible, early latent, late latent, infectious, and recovered. These compartments are replicated across four population strata, which individuals traverse via incarceration and release: never incarcerated, currently incarcerated, recent history of incarceration, and distant history of incarceration. We distinguish between recent and distant incarceration history to account for the elevated risk of recidivism, tuberculosis, and mortality in the early period post-release^6,7,14^. The model does not include HIV, drug resistance, gender/sex, or age structure and excludes children aged 14 years and under who are assumed to not be at risk of incarceration. We assume that higher tuberculosis risk in prisons results from higher effective contact rates, higher disease progression rates, and lower diagnosis rates compared to outside prison. We include low levels of mixing between incarcerated and non-incarcerated individuals to represent interactions with prison staff and visitors.

We fit the model independently for each country to incarceration and tuberculosis data from 1990 to 2023. Yearly calibration targets included incarceration prevalence, prison entries (admissions), and recidivism, as well as total and within-prison tuberculosis incidence and notification rates (**Tables S1-S2**). We accounted for uncertainty by sampling from distributions for calibration targets and for a subset of parameters that were fixed during calibration (**Tables S3-S4**). For each sample of calibration targets and fixed parameters, we then ran optimization algorithms to calibrate the remaining parameters, obtaining at least 1000 fitted parameter sets per country **(Figure S4**).

For time-varying parameters, we let the model reach equilibrium with baseline values and then applied rates of change starting in the year 1990. Changes in incarceration prevalence over time were achieved through changes in prison entry and release rates; changes in tuberculosis incidence and notification rates were achieved through changes in effective contact rates and diagnosis rates (**Table S5, Figure S5**). We also accounted for COVID-19 pandemic-related changes (**Table S6**).

Details on model specification, assumptions, calibration, and validation are provided in the Appendix.

### Excess burden estimates

For each country, we quantified the excess population-level tuberculosis incidence attributable to the rise in incarceration prevalence since 1990 by simulating a counterfactual scenario where incarceration prevalence and dynamics remained stable at 1990 levels. To operationalize this, for each set of fitted parameters, we re-ran the model from 1990, with time-dependent changes in prison entry and release rates turned off. We also eliminated time-dependent changes in the effective contact rate within prison, which we assumed to be linked to growing prison populations. We then calculated the excess burden as the relative and absolute difference in population tuberculosis incidence between the observed and counterfactual scenarios. We report excess burden estimates in 2019 and 2022 but use 2019 for our main estimates due to COVID-19-related uncertainty. We also analyze where excess incident cases arose—i.e., where individuals progressed or relapsed to infectious tuberculosis disease—and where cases were diagnosed and notified.

### Sensitivity analyses and meta-modeling

We performed five sensitivity analyses varying key assumptions around natural history, differences across strata, changes over time, and mixing. We additionally conducted linear regression meta-modeling using a multi-level model to identify parameters associated with variation in excess burden estimates^15^. The **Appendix** includes details on sensitivity analyses and meta-modeling.

### Transmission population attributable fraction (tPAF)

To estimate the tPAF for incarceration among individuals aged 15 and older, we simulated a scenario where incarceration prevalence was gradually reduced to zero by 2009 (**Appendix**). After ten years of no new exposure to incarceration, we calculated the tPAF for incident cases in 2019 as follows^16^:

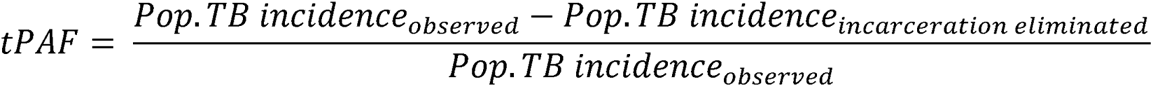

We compared our estimates of the tPAF for incarceration with WHO’s country-specific estimates of the fraction of all incident cases attributable to each of five major tuberculosis risk factors in 2019. We note that risk factors may be overlapping, and that WHO’s estimates apply to varying age groups: undernutrition, all ages; HIV, all ages; alcohol use disorders, age ≥15; smoking, age ≥15; diabetes, age ≥18. For diabetes, the PAF is reported as a fraction of all cases among individuals age ≥15, rather than a true PAF, and therefore may be an overestimate.

### Future policy scenarios

We simulated various incarceration scenarios over a ten-year period (2024-2034) and estimated their impacts on future population tuberculosis incidence. Under the reference or “stable” scenario, prison entry rates and average duration of incarceration remain constant. Under the “continue trends” scenario, entry rates and duration undergo the same relative net change between 2024 and 2034 as over the prior ten years. The decarceration scenarios involve gradual 25% or 50% reductions in entry rates, duration, or both by 2034. We computed the percent difference in projected population tuberculosis incidence in 2034 under each scenario compared to that expected under the stable scenario.

In El Salvador, the prison population has nearly tripled since March 2022 under a continued state of emergency^17^. We estimate the excess population TB incidence in 2024 attributable to the recent state of emergency by simulating a counterfactual scenario without the observed rise in incarceration prevalence since March 2022. We additionally simulated the following future scenarios from 2024-2034: 1) continuation of current entry and release rates under the state of emergency; 2) passive abatement through entry and release rates gradually returning to their pre-emergency levels by 2034; and 3-5) active cessation of the state of emergency by 2025 and reversion of incarceration prevalence to its approximate pre-emergency level in ten, five, or two years (i.e., by 2034, 2029, or 2026, respectively), with continued decarceration thereafter. Reversion of incarceration prevalence to pre-emergency levels under scenarios 3-5 is achieved through entry and release rates changing promptly by 2025. Rather than comparing to a reference scenario, we computed the percent change in population tuberculosis incidence in 2034 under each scenario compared to 2021. Methods for future projections are detailed in the **Appendix**.

## RESULTS

Argentina, Brazil, Colombia, El Salvador, Mexico, and Peru exhibited wide variability in the population-wide and within-prison burden of tuberculosis between 1990 and 2019 **(Table 1)**. In 2019, the tuberculosis notification rate in prisons was a median 28.7 (IQR 13.1-31.6) times the population-wide notification rate (**Table 1**). Calibrated effective contact rates, disease progression rates, and diagnosis rates in prison were a median 6.8 (IQR 2.5-11.9), 2.3 (2.0-2.9), and 0.55 (0.44-0.59) times those in the community, respectively (**Table S7**).

**Table 1.**
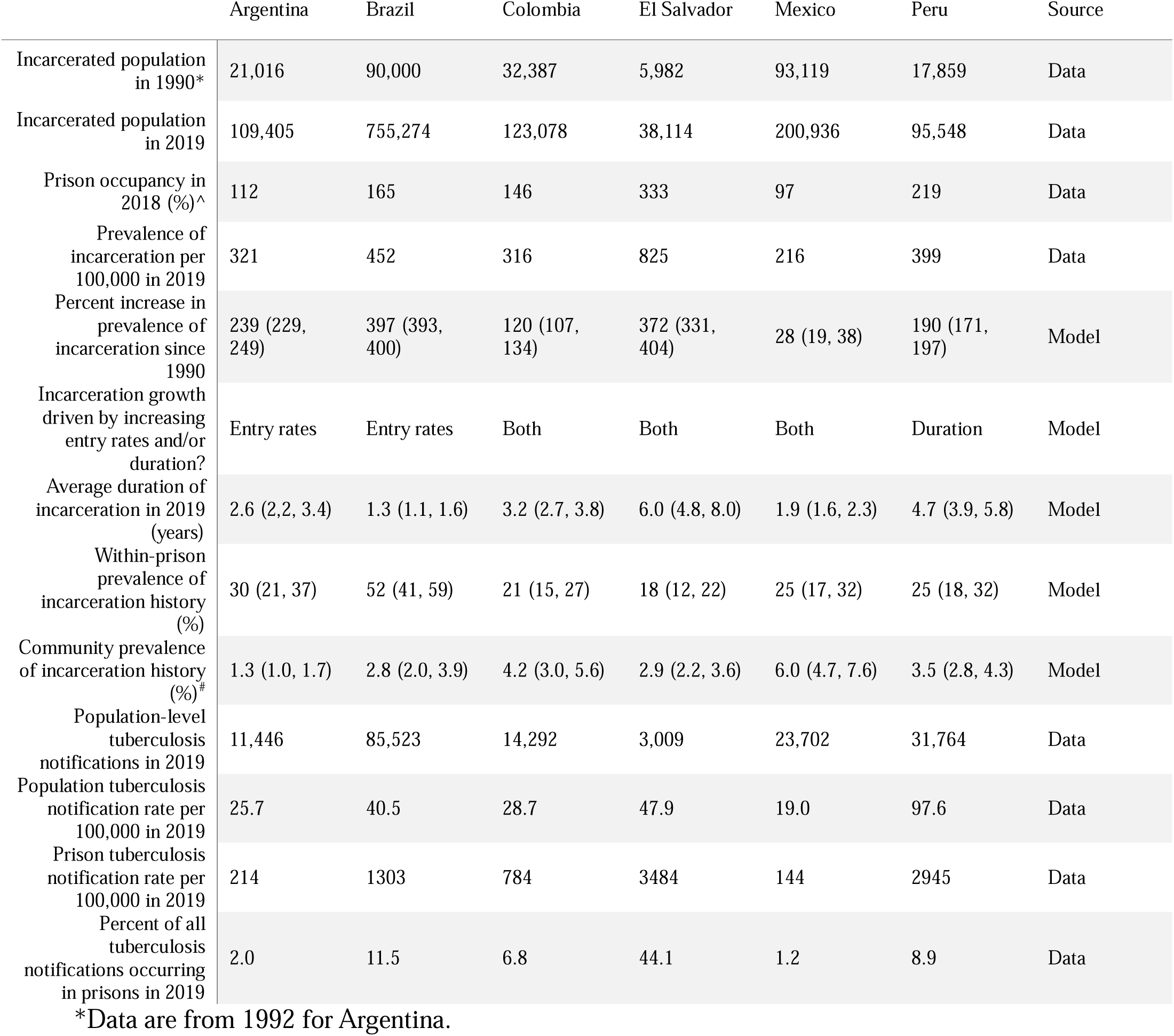
Incarceration- and tuberculosis-related characteristics by country. All population-wide prevalence estimates are for the population aged 15 and older. Data sources are detailed in Tables S1-S2 of the Appendix. For model outputs, medians are shown with 95% uncertainty intervals in parentheses.

Between 1990 and 2019, the prevalence of incarceration among the population aged 15 and older more than doubled in all countries except Mexico, reaching a median 360 (IQR 317 to 439) per 100,000 in 2019 across the six countries (**Table 1**). This historical rise was driven by an increase in prison entry rates (Argentina and Brazil), an increase in average duration of incarceration (Peru), or both (El Salvador, Colombia, Mexico) (**Table S8**). By 2019, the average duration of incarceration ranged from 1.3 years (95% UI 1.1-1.6) in Brazil to 6.0 years (4.8-8.0) in El Salvador (**Table 1**. Further, the percent of the prison population with prior incarceration history ranged from 18% (95% UI 12-22) in El Salvador to 52% (41-59) in Brazil. Such differences in incarceration dynamics contribute to the heterogeneity in community prevalence of incarceration history among the population age 15 and older, which ranged from 1.3% (95% UI 1.0-1.7) in Argentina to 6.0% (4.7-7.6) in Mexico (**Table 1**). Across all six countries in 2019, while 1.3 million people were incarcerated at any given time, we estimate that an additional 13.4 million (95% UI 11.4-15.7 million) people were living with incarceration history.

Compared to a counterfactual scenario where incarceration prevalence remained constant since 1990, the observed rise in incarceration prevalence since 1990 resulted in an estimated 34,393 (95% UI, 28,295-42,579) excess incident cases in 2019 across the six countries (**Figure 2A-B**, **Table 2**). The excess population tuberculosis incidence in 2019 varied widely across countries, ranging from 6% or 1.3 (95% UI 0.8-2.1) cases per 100,000 person-years in Mexico to 134% or 32.2 (25.5-41.3) cases per 100,000 person-years in El Salvador (**Table 2**). Estimates for the year 2022 were comparable (**Table S9**). Sensitivity analyses varying several assumptions did not substantively change our results (**Figure S9**).

**Figure 2.**
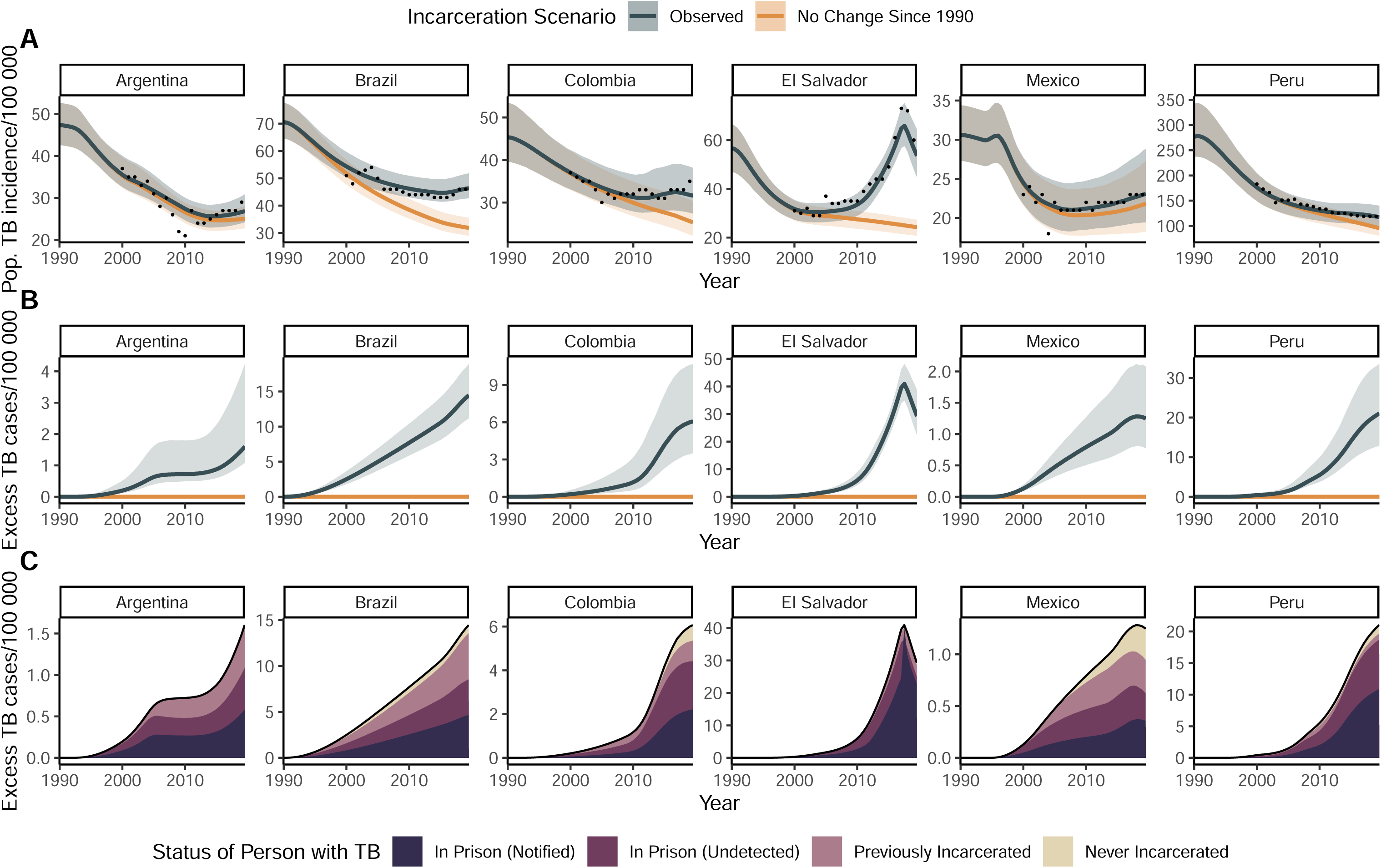
Excess population tuberculosis incidence attributable to the rise in incarceration prevalence since 1990. A) Population tuberculosis incidence per 100,000 person-years under the observed and counterfactual (no rise in incarceration since 1990) scenarios. Black points represent population tuberculosis (TB) incidence estimates from the World Health Organization, which are available beginning in 2000. Solid lines and shaded bands represent the median and 95% UI respectively. B) Excess population-wide incident tuberculosis cases per 100,000 person-years. C) Median estimates of excess cases, stratified by population subgroup in which they occurred, and for incident cases occurring in prison, additionally stratified by whether the disease was notified or undetected during incarceration. All model results are for the population age 15 and older.

**Table 2.** Estimates of population tuberculosis incidence attributable to incarceration in 2019. All estimates are at the population-level among individuals aged 15 and older. Incidence rate ratios and excess burden estimates were obtained from comparing incident tuberculosis cases in 2019 between the observed scenario of the historical rise in incarceration and the counterfactual scenario of no change in incarceration prevalence since 1990. The transmission population attributable fraction in 2019 was estimated using a scenario where incarceration prevalence was reduced to zero by 2009. 95% uncertainty intervals are shown in parentheses.

|  | Incidence rate ratio for observed vs. counterfactual | Excess cases per 100,000 person-years relative to counterfactual | Absolute excess cases relative to counterfactual | Transmission population attributable fraction (%) |
| --- | --- | --- | --- | --- |
| Argentina | 1.06 (1.04, 1.16) | 1.5 (1.0, 3.9) | 506 (344, 1344) | 8.4 (6.0, 18.6) |
| Brazil | 1.44 (1.32, 1.59) | 14.1 (10.9, 18.4) | 23497 (18160, 30739) | 36.9 (29.5, 45.1) |
| Colombia | 1.23 (1.13, 1.42) | 6.0 (3.4, 10.6) | 2337 (1338, 4135) | 21.8 (14.1, 34.7) |
| El Salvador | 2.34 (2.03, 2.69) | 32.2 (25.5, 41.3) | 1489 (1178, 1907) | 58.1 (51.6, 64.1) |
| Mexico | 1.06 (1.04, 1.09) | 1.3 (0.8, 2.1) | 1180 (740, 1957) | 7.5 (4.8, 11.6) |
| Peru | 1.21 (1.13, 1.34) | 20.6 (12.6, 33.0) | 4922 (3028, 7899) | 23.3 (16.7, 34.4) |

The burden of excess incident cases that arose (i.e., progressed to disease or relapsed) in prisons in 2019 exceeded that of excess cases *diagnosed* within prisons by 81%, ranging from an additional 10% in El Salvador to 102% in Colombia **(Figure 2C)**. Furthermore, a considerable fraction of the excess burden in 2019 was comprised of incident cases arising among formerly incarcerated individuals, particularly in countries with shorter average duration of incarceration **(Figure 2C)**. For instance, the percent of excess cases arising in the community among formerly incarcerated individuals was 34% (95% UI 24-45) in Argentina, 34% (26-42) in Brazil, and 26% (16-36) Mexico (**Table S11)**. In all countries, estimated tuberculosis incidence rates among individuals with recent or incarceration history were much higher than population-wide incidence rates (**Figure S10, Table S12**).

Collectively across countries, incarceration was the leading determinant compared to other key tuberculosis risk factors, accounting for an estimated 27.2% (95% UI, 20.9-35.8) of incident cases in 2019 among the population aged 15 and older. The country-specific tPAF of incarceration in 2019 reached 58.1% (95% UI, 51.6-64.1) in El Salvador, 36.9% (29.5-45.1) in Brazil, 23.3% (16.7-34.4) in Peru, 21.8% (14.1-34.7) in Colombia, 8.4% (6.0-18.6) in Argentina, and 7.5% (4.8-11.6) in Mexico (**Table 2**). Despite this variability, the country-specific tPAF for incarceration was consistently greater than or commensurate with PAFs for other major risk factors (**Figure 3)**. Moreover, our median tPAF estimate was 1.3 to 6.3 times the percent of all tuberculosis notifications occurring in prisons in 2019 (**Figure 3)**.

**Figure 3.**
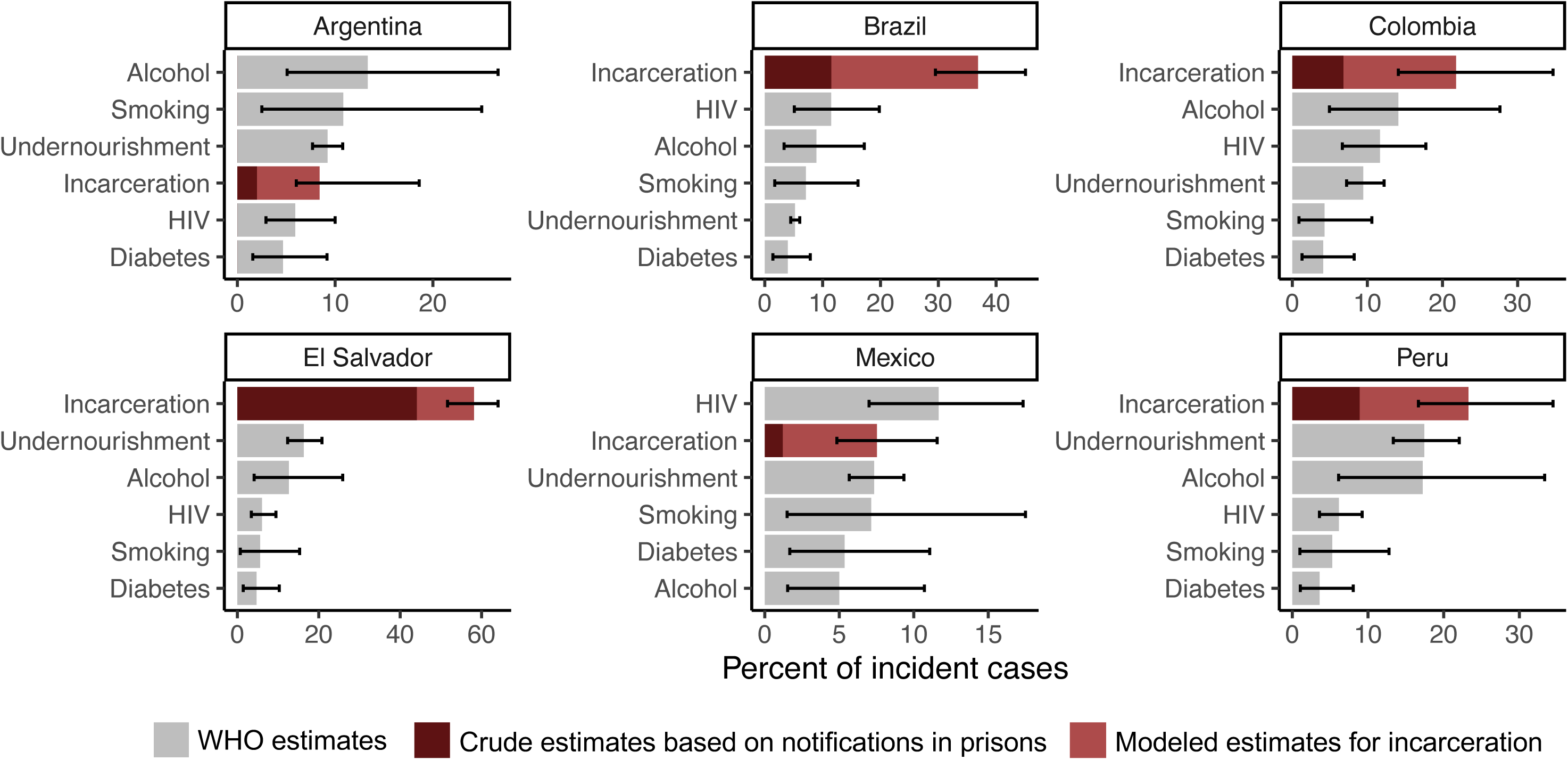
Population attributable fraction (PAF) for incarceration and other tuberculosis risk factors. Median estimates and uncertainty intervals for the percent of population-level incident tuberculosis cases in 2019 that can be attributed to each risk factor. The “crude” estimate of the population attributable fraction for incarceration is based on the percent of all notified tuberculosis cases occurring in prisons. Estimates for all other risk factors are from the World Health Organization (WHO). Risk factors are listed in descending order by PAF for each country. Estimates correspond to different age groups: incarceration, age >15; undernutrition, all ages; HIV, all ages; alcohol, age >15; smoking, age >15; diabetes, age >18 (see **Methods**).

We projected the impact of future incarceration policies, implemented from 2024 to 2034, on population tuberculosis incidence in 2034. Future projections for El Salvador are described below. For all other countries, projected incarceration prevalence in 2034 under each scenario is shown in **Figure 4A** and **Table S13.** If recent incarceration trends continue, projected population tuberculosis incidence in 2034 would be slightly (<3%) higher in Peru, Argentina, and Mexico, and slightly lower in Colombia and Brazil **(Figure 4B)**. More active decarceration interventions—for instance, a 50% decrease in prison entry rates and duration of incarceration— could reduce population tuberculosis incidence in 2034 by an estimated 28.9% (95% UI 22.0-36.7) in Brazil, 16.4% (11.4-23.3) in Peru, 13.7% (8.9-21.3) in Colombia, 10.3% (7.1-16.9) in Argentina, and 3.0% (1.3-5.7) in Mexico.

**Figure 4.**
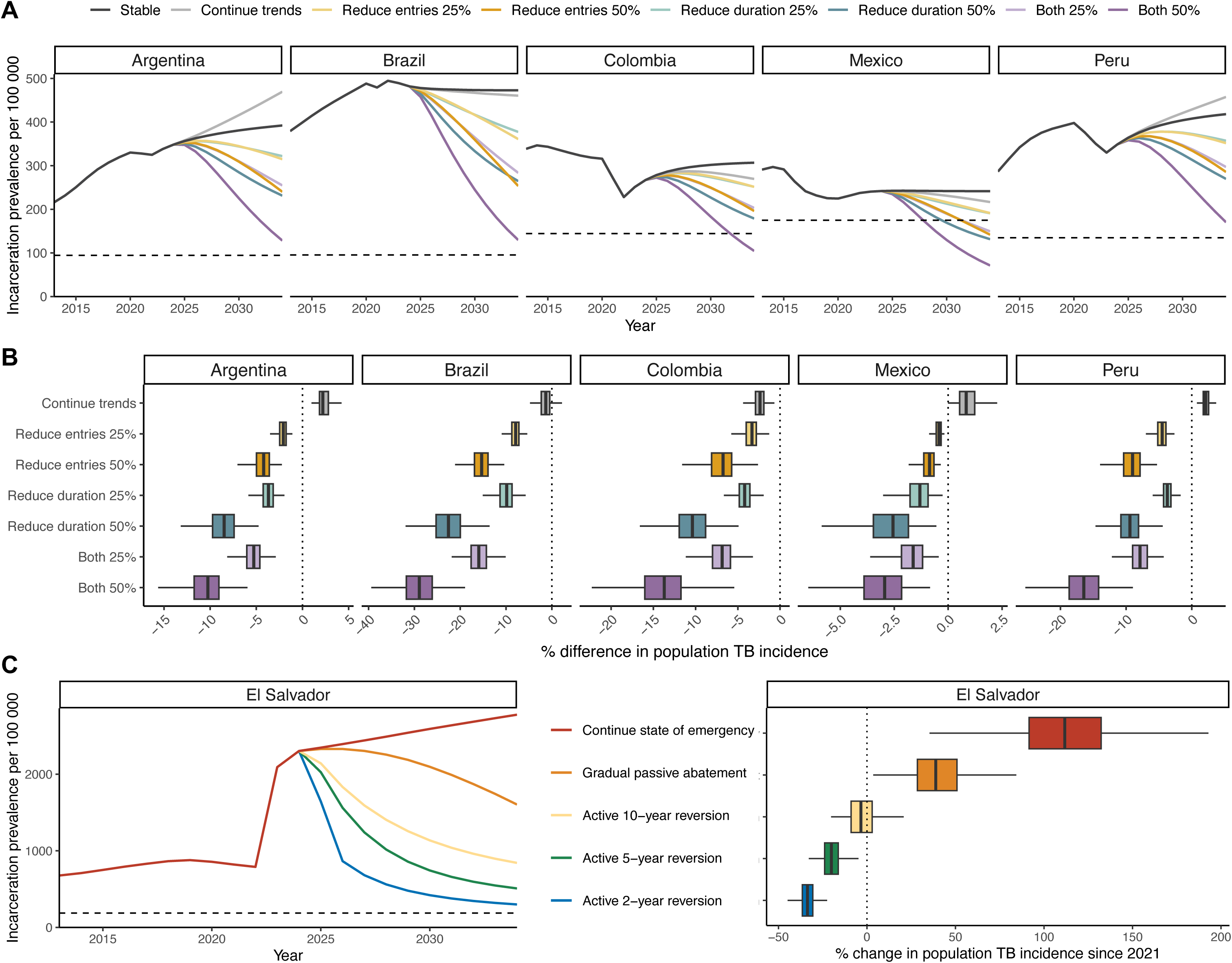
Projected impacts of incarceration-related interventions on future population tuberculosis incidence. A) Median incarceration prevalence per 100,000 population aged 15 and older under incarceration scenarios implemented between 2024 and 2034: stable entry and release rates (reference scenario), continuation of trends from prior ten years, and 25% or 50% reduction in prison entry rates, duration of incarceration, or both by 2034. The dashed horizontal line represents incarceration prevalence in 1990. B) Percent difference in population tuberculosis incidence in 2034 under each incarceration scenario, relative to the reference scenario of stable entry and release rates. Outliers are not shown. C) Left: Median incarceration prevalence under each incarceration scenario in El Salvador: continuation of entry and release rates under the state of emergency, passive abatement through gradual reversion of entry and release rates to pre-emergency levels by 2034, active cessation and approximate restoration of pre-emergency incarceration prevalence in ten years, or restoration of pre-emergency prevalence in five years or two years with continued decarceration thereafter. The dashed horizontal line represents incarceration prevalence in 1990. Right: Percent change in population TB incidence since 2021 under each scenario.

In El Salvador, we project that population TB incidence in 2024 is 2.1 (95% UI 1.8-2.4) times as high as expected without the recent state of emergency, corresponding to 2,444 (95% UI 1,562-3,245) excess incident cases in 2024 (**Figure S11**). Maintaining the state of emergency for ten years is projected to increase population tuberculosis incidence in 2034 by 112% (95% UI 63-176) compared to pre-emergency in 2021 **(Figure 4C)**. A gradual, passive abatement of the state of emergency would still increase population tuberculosis incidence in 2034 by a projected 39% (95% UI 13-72). In contrast, prompt and active cessation of the state of emergency and reversion of incarceration prevalence to approximate pre-emergency levels by 2034 could restore population tuberculosis incidence in 2034 to its approximate rate in 2021. More decisive actions to revert pre-emergency incarceration prevalence in two or five years and continue further decarceration thereafter could reduce population tuberculosis incidence in 2034 by as much as 34% (95% UI 25-42) compared to 2021.

## DISCUSSION

Across six Latin American countries, more than 34,000 incident cases in 2019 can be attributed to the rise in incarceration since 1990. Collectively in these countries, incarceration accounts for an estimated 27.2% (95% UI, 20.9-35.8) of incident cases in 2019 among individuals aged 15 and older, a far greater fraction than any other determinant. Against the backdrop of the region’s alarming increase in tuberculosis incidence over the last decade, we project that policies to reduce incarceration prevalence may considerably reduce future population tuberculosis incidence. Together, our results implicate incarceration as the leading population-level driver of the tuberculosis epidemic in Latin America. In addition to improving prison conditions and implementing biomedical interventions in prisons, criminal legal reforms and development of non-carceral alternatives will be critical to re-ignite progress towards tuberculosis elimination.

Our results elucidate the harmful impacts of decades of punitive policies on the tuberculosis epidemic in the region. Beginning in the 1990s, amidst rising crime and public support for “tough-on-crime” initiatives, governments in Latin America expanded police and prosecutorial activity, criminalized new acts, and imposed harsher sentences, including for minor offenses^18,19^. Consequently, prison populations surged, largely comprised of individuals detained for property- and drug-related offenses^18,20^. Meanwhile, inadequate investments in the penitentiary system led to severe overcrowding, inhumane living conditions, deficient health care, corruption among staff, uprisings, and self-governance^18,21^. Today, the incarceration rate in Latin America is twice the global rate and higher than all other regions except North America^2^. Criminologists argue that prisons have been ineffective and even counterproductive in curbing crime in the region^18,21^. Instead, they have created new challenges, including a worsening crisis of tuberculosis in prisons.

In the present study, we show that the scope and magnitude of this crisis is even larger than previously recognized. To date, research and policy guidance has focused on tuberculosis occurring within prisons^3,22–25^. However, unlike other TB risk factors, incarceration is highly dynamic. The constant flow of people who are newly incarcerated and released yields a much larger population ever exposed to the high-risk carceral environment, which we estimate across the six countries is over 11 times the size of the population in prison at any given time. By accounting for this phenomenon and its interplay with the variable latent period of tuberculosis, we obtained attributable burden estimates that far exceeded crude, static estimates based on notifications in prisons. Of note, most of the difference was due to under-detection in prisons and progression to disease following release, rather than onward transmission. Thus, while traditional PAF estimates for other TB determinants may also be underestimated due to not accounting for onward transmission, incarceration is particularly subject to under-recognition by conventional approaches that do not account for its dynamic nature. Policy guidance and future research should recognize incarceration as a tuberculosis driver and social determinant with effects that transcend prison walls.

We also demonstrate the potential impact of alternative incarceration policies on the tuberculosis epidemic in the region. For instance, policies that decrease prison admissions and duration by 50% could reduce future population tuberculosis incidence by more than 10% in Brazil, Peru, Colombia, and Argentina, countries which encompass the vast majority of the region’s tuberculosis burden. In El Salvador, which already had an exorbitant tPAF for incarceration prior to 2022, the state of emergency is projected to have catastrophic consequences for tuberculosis. We predict that swift, resolute termination of the state of emergency could enable a return to pre-emergency incidence by 2034, and that further decarceration can recover, at least in part, a decade of lost opportunity for tuberculosis progress. Such measures have precedent in Kazakhstan, where KNCV and Penal Reform International co-led comprehensive efforts to address tuberculosis in prisons, integrating biomedical interventions with decriminalization reforms, implementation of alternatives to incarceration, and improvements in prison conditions^26^. Following expansion of the program in 2000, incarceration prevalence decreased by 70% and with it, the rate of tuberculosis in prisons by 90%^27^. Therefore, decarceration interventions, especially if coupled with biomedical interventions and efforts to improve prison conditions, have substantial potential to accelerate progress towards 2035 End TB strategy targets.

Our estimates of the tuberculosis burden attributable to incarceration vary greatly across the six countries included, correlating with incarceration prevalence and country-specific disparities in tuberculosis risk between prisons and the general population. Between-country variation in where excess cases arise can also be attributed to distinct carceral dynamics across countries. For instance, in countries with a longer average duration of incarceration, like El Salvador and Peru, our model predicts that the vast majority of excess incident cases occur within prisons^5^. Conversely, in countries with a shorter average duration of incarceration, i.e., Brazil and Mexico, a greater proportion of excess incident cases occur in the community after prison release. Therefore, it is crucial to consider incarceration dynamics and changing carceral policies in identifying optimal intervention strategies. These insights may generalize to other countries and regions. Specifically, in most other settings with lower incarceration rates and less disparity in tuberculosis rates between prisons and the general population, incarceration may play a lesser role in the tuberculosis epidemic. Nonetheless, in all settings, the true incarceration-attributable tuberculosis burden likely exceeds crude estimates based on tuberculosis occurring within prisons, especially where prison turnover rates are high.

In response to this public health crisis, bold and decisive investments and actions are needed. First, international health agencies and national tuberculosis programs must improve reporting of incarceration as a structural determinant of tuberculosis. This includes collecting information on incarceration history in case notifications databases and including current and past incarceration as a key risk factor in WHO’s Global Tuberculosis Report^8^. Given the stigma and discrimination faced by individuals with incarceration history, procedures for collecting this information in a sensitive manner should be developed alongside stakeholders with lived experience of incarceration^28^. Second, effective strategies to prevent, detect, and treat tuberculosis in incarcerated and formerly incarcerated individuals must be identified, incorporated in national guidelines, and implemented at scale^29,30^. While existing research has focused on prison-based interventions, future work should expand to include formerly incarcerated individuals and their community contacts.

Lastly, and equally as important, governments must implement structural reforms to reduce the prison population. While our study focused on tuberculosis, incarceration exposure has been linked to other adverse health outcomes^14,31,32^. Therefore, decarceration strategies, especially in conjunction with efforts to transform conditions of confinement, have the potential to both accelerate tuberculosis progress and improve population health at large. Currently, political will and public support for such measures remain low. However, calls are growing for governments to improve prison conditions, decriminalize minor offenses, reduce pre-trial detention, and develop restorative justice-based alternatives to incarceration, with several initiatives underway across the region^18,20,21,33–36^.

Our study has several limitations. First, for four of six countries—all except Brazil and Colombia—empirical prison-based active case-finding studies were unavailable, so prison incidence estimates for model calibration were based on a regional case detection ratio. For these countries, findings should be viewed as estimates within a plausible range of uncertainty. Second, deterministic compartmental models are unable to capture the full range of complexity in real-world phenomena. The extent to which we were able to incorporate complexity in our model was constrained by inadequate data to inform model parameters and assumptions. For instance, our model did not account for age, gender/sex, socioeconomic status, HIV, heterogeneity in duration of incarceration, heterogeneity in infectiousness, or MDR-TB, which is less common in prisons in the Americas than in other regions^37^. Accounting for HIV or MDR-TB, which may be exacerbated in prisons, may increase estimates of the incarceration-attributable burden^12^. Accounting for other factors like age or socioeconomic inequities that affect mixing and tuberculosis risk may result in lower estimates for incarceration^24^. Moreover, we had little to no data to inform mixing assumptions or stratum-specific parameters for formerly incarcerated individuals. In these cases of insufficient data, we used wide parameter uncertainty distributions and varied our assumptions in sensitivity analyses, with our findings generally remaining robust. However, the dearth of reliable, publicly accessible data on incarceration and tuberculosis must be urgently addressed.

Next, our future projections are subject to great uncertainty, including uncertainty around how the COVID-19 pandemic has affected and will continue to affect tuberculosis and incarceration. We were unable to model specific policies or reforms (i.e., decriminalization of drug use) due to insufficient data. Our future simulations also do not include changes in any other dimension aside from prison entry and release rates, such as improvements in prison conditions or scale-up of biomedical interventions. Generally, our historical counterfactual and future policy simulations are simplistic, modifying incarceration in isolation from what is inevitably an intricate web of upstream and downstream social, economic, political, and institutional forces that themselves also affect population health and tuberculosis. Nonetheless, our findings underscore the substantial potential for criminal legal reforms to reduce tuberculosis burden in Latin America, impacts which could be enhanced by additional prison- and community-based interventions.

To date we have failed to appreciate the full extent to which rising incarceration has undermined tuberculosis control in Latin America. Our estimates of the outsized tuberculosis burden attributable to incarceration eclipse those of other determinants that currently receive far greater attention. However, this exceptional excess burden must not be regarded as inevitable. Health agencies, national tuberculosis programs, ministries of justice, and other key stakeholders should undertake bold commitments and actions to elevate the prominence of incarceration in national and international strategies for tuberculosis control and elimination, accounting for impacts beyond prison walls. These strategies should take an integrated health and human rights approach, combining biomedical interventions and improvements in prison conditions with actions to enable decarceration. Such measures will be critical to advancing towards regional and global tuberculosis elimination targets.

## Data Availability

Data and code for the present study have been deposited in the GitHub respository, tb_incarc_mod.

https://github.com/yemloo/tb_incarc_mod

## Author contributions

YEL and JRA conceived and designed the study. YEL and YM collected data with assistance from LML. YEL and YM developed and calibrated the incarceration sub-model. YEL developed and calibrated the primary tuberculosis model, performed analyses, and made tables and figures. SC, JDGF, and JRA assisted with model development and calibration. JRA, JC, JDGF, and TC provided guidance on analyses. YEL wrote the first draft of the manuscript. YM and MB contributed to writing of subsequent drafts. All authors contributed to data interpretation and critical revision of the manuscript. YEL and YM accessed and verified the underlying data. All authors approved the final version of the manuscript and agreed to submission.

## Acknowledgments

This study was funded by the National Institutes of Health (grant numbers 5R01AI130058 and 5R01AI149620). YEL is funded by the Stanford Knight Hennessy Scholars Program, the National Science Foundation Graduate Research Fellowship, and the Gerald J. Lieberman Fellowship from the Stanford Office of the Vice Provost for Graduate Education. We thank Edwin Segura, Noah Bullock, Hernán Olaeta, the National Penitentiary and Prison Institute of Colombia, and Victor Peña Garcia for providing data and useful insights.

## Data sharing

No individual participant data were collected in this study. Data and code used for modeling are available at github.com/yemloo/tb_incarc_mod.

