## Supplementary Appendix for "Mass incarceration as a driver of the tuberculosis epidemic in Latin America and projected impacts of policy alternatives: A mathematical modeling study"

**Table of Contents**

Methodological Details

1. Model structure, assumptions, and equations

2. Parameter calibration and uncertainty analysis

3. Model validation

4. Historical counterfactual scenarios

5. Sensitivity analyses and meta-modeling

6. Future projections

Table S1. Data availability and sources for incarceration-related calibration targets.

Table S2. Data availability and sources for tuberculosis-related calibration targets.

Table S3. Description of model parameters.

Table S4. Uncertainty distributions for calibration targets.

Table S5. Intervals and direction of change in time-varying parameters.

Table S6. Changes in model parameters during COVID-19 pandemic.

Table S7. Relative values of calibrated within-prison tuberculosis parameters in 2019.

Table S8. Modeled percent change in prison entry and release rates from 1990-2019.

Table S9. Estimates of excess population tuberculosis incidence attributable to mass incarceration in 2022.

Table S10. Results of multi-level meta-modeling.

Table S11. Percent of excess incident cases in 2019 among formerly incarcerated individuals.

Table S12. Model outputs for stratum-specific tuberculosis incidence rates per 100,000 person-years in 2019.

Table S13. Incarceration prevalence in 2034 under future incarceration scenarios.

Table S14. Additional parameters to model El Salvador’s state of emergency (SoE).

Table S15. Incarceration prevalence in 2034 under future incarceration scenarios in El Salvador.

Figure S1. Schematic of meta-population dynamic compartmental model.

Figure S2. Posterior distribution for regional case detection ratio (CDR) in prisons.

Figure S3. Schematic of simple population transition model for calibrating incarceration-related parameters.

Figure S4. Prior and posterior distributions for calibrated model parameters.

Figure S5. Time-varying parameters.

Figure S6. Model fit to incarceration-related calibration targets.

Figure S7. Model fit to tuberculosis-related calibration targets.

Figure S8. Model consistency with validation data.

Figure S9. Results from five sensitivity analyses.

Figure S10. Stratum-specific tuberculosis incidence rates.

Figure S11. Projected impacts of El Salvador’s state of emergency on population TB incidence.

References

**Methodological Details**

1. **Model structure, assumptions, and equations**

We developed a deterministic, meta-population dynamic compartmental transmission model to describe incarceration and tuberculosis dynamics **(Figure S1)**. We used a simple tuberculosis natural history structure that does not include age, HIV, or drug resistance. The model accounts for observed and projected population growth using data from World Population Prospects. The model is described by a set of ordinary differential equations implemented using *ode* in R. Equations are provided below and parameters are described in **Table S3.**

In the tuberculosis dimension of the model, susceptible individuals (S) who are infected develop early latent infection (E), after which they can progress directly to infectious, active disease (I) or transition to late latent (L), where they experience a lower rate of progression to active disease. Those with active disease can enter the recovered state (R) through self-cure or diagnosis and treatment, a transition which occurs instantaneously. Recovered individuals can relapse to active disease. Individuals in both late latent and recovered states can be re-infected to the early latent state at a reduced rate compared to susceptible individuals.

All tuberculosis states are reproduced across four incarceration-related population strata, indicated by lowercase subscripts in the model schematic and equations. Individuals enter the model in the **n**ever incarcerated stratum (n) and transition to the **p**rison stratum (p) upon incarceration. Upon release from prison, individuals transition to the **r**ecent history of incarceration stratum (r). Those who remain out of prison transition over time to the **d**istant history of incarceration stratum (d). We assume higher risk of re-incarceration and mortality among those with recent but not distant incarceration history, compared to never incarcerated individuals. This is based on studies showing increased risks of recidivism and mortality in the early period post-release that attenuate over time^1-3^.

We assume individuals are born into the model at age 15 in susceptible, early latent, or late latent states. The proportion entering the model as susceptible is based on the cumulative risk of infection by age 15, expressed as $1-e^{-\left( \beta_{cc}\frac{I_{c}}{N_{c}}\frac{I_{n}}{N_{n}} +\beta_{cp}\frac{I_{p}}{N_{p}} \right)\left( 15 \right)}$. Among those entering the model with latent infection, we assume a 1:6 ratio of early to late latent infection based on evidence showing that most individuals who progress to disease do so within two years of infection. We also test a 1:2 ratio, based on an assumption that risk of disease progression remains elevated for the first five years of infection, in a sensitivity analysis (section 5).

We assume that the elevated rates of tuberculosis in prisons are the result of a higher effective contact rate, higher fast progression rate, and a lower diagnosis rate. These parameters are calibrated to incidence and notifications data (see section 2). Our assumptions are supported by the following rationale and evidence:

1) Prisons are dense congregate settings, which is aggravated by severe overcrowding and poor ventilation. This results in incarcerated individuals having more contacts per unit time, and higher probability of infection given contact^4^.
2) Studies have shown that longer duration and higher “dose” of exposure are associated with higher risk of disease progression^5,6^. Infections acquired in prisons likely arise from exposures of higher dose and frequency due to overcrowding, poor ventilation, and elevated disease prevalence^4^. Incarcerated individuals may also experience malnutrition, inadequate healthcare, psychological distress, social isolation, and violence, as supported by surveys conducted in prisons throughout the region^7^. All of these may worsen health and result in increased risk of disease progression^8^.
3) Case detection rates are estimated to be lower in prison than in the general population based on a recent modeling study integrating notifications data with incidence and prevalence studies in prisons^9^.

We also assume that the recent incarceration history stratum has an elevated disease progression rate and a reduced diagnosis rate, for the following reasons:

1) Individuals recently released from prison may face stigma and numerous barriers to obtaining stable housing, employment, healthcare access, nutrition, and other basic needs^8,10^. These factors may contribute to lower case detection rates and increased risk of disease progression (through worsened health).
2) Individuals recently released from prison may also experience lasting negative impacts of incarceration (i.e. malnutrition, inadequate healthcare, stress, isolation) on health^8,10^. Additionally, recently released individuals likely acquired infection through high-dose exposure in prison (i.e., due to overcrowding, poor ventilation, elevated prevalence), this may result in an increased risk of disease progression even after prison release^4-6^.
3) Data linkage studies from Brazil and Paraguay have shown that tuberculosis notification rates are higher for up to 7-8 years following release^11,12^. This long period of elevated risk is likely due not only to undiagnosed tuberculosis upon release but also to new progression to disease after release.

To operationalize this, fast progression rates and diagnosis rates in the **r** stratum are calculated as weighted averages of their respective values in prison and in the other two non-incarcerated strata (**n** and **d**). The weights are sampled from uniform distributions ranging from 0 to 1, where 1 results in the same value as in prison, and 0 results in the same value as in the **n** and **d** strata. We also include a sensitivity analysis where progression and diagnosis rates in the **r** stratum are equivalent to those in the **n** and **d** strata (section 5).

To fit to observed changes in incarceration prevalence over time, we allowed prison entry and release rates to change, with periods and magnitude of change informed by data on admissions and releases over time **(Table S1)**. For periods where admissions and/or release data were not directly available, decisions around changes over time were informed by the following:

1) Data on sentence lengths over time from national penitentiary departments or surveys. These data cannot be used directly as many individuals do not serve the entirety of their sentence in prison; moreover, they are subject to biases inherent to cross-sectional data. However, assuming that the proportion of sentence served remain relatively stable, these data can provide some insight into how average duration of incarceration has changed over time.
2) Data on the proportion of the prison population in pre-trial detention, who generally spend less time incarcerated than individuals who are convicted.
3) Legal reforms that were likely to affect admissions and releases: for instance, creation of new crimes is likely to increase admission rates, while increases in minimum or maximum sentences are likely to decrease release rates.

Data on average duration of incarceration were not available, but this parameter can be triangulated using data on the prevalent population, inflows, and outflows. Modeled estimates of the average duration of incarceration in **Table 1** were calculated as $\frac{1}{r+{mu}_{p}}$, where $r$ and ${mu}_{p}$ represent modeled prison release and mortality rates, respectively.

To fit to observed changes in population-wide tuberculosis incidence and notification rates over time, we allowed the effective contact rate and diagnosis rate to change over time outside prison. To fit to observed changes in prison tuberculosis incidence and notification rates, we allowed the effective contact rate and the diagnosis rate in prison to change over time. We include a sensitivity analysis where the effective contact rate in prison does not change over time (section 5).

We operationalized time-varying parameters using baseline values and either rates of change or multipliers, some of which were calibrated and others sampled from uncertainty distributions. We brought the model to equilibrium as governed by baseline parameters before temporal changes to parameter values began in 1990 or later.

In the main analysis, we assume that all three population strata outside prison, which we simplify as “community” (c), mix proportionately with equal effective contact rates. We therefore use the same effective contact rate ($\beta_{cc}$) for all intra- and inter-stratum mixing outside prison. We implement assortative mixing outside prison in a sensitivity analysis (section 5).

In the main analysis, to account for possible transmission between incarcerated people and non-incarcerated prison staff or visitors, we allow low levels of mixing between people in prison and people outside prison. Prior genomic epidemiologic studies in Brazil provide evidence for direct transmission between incarcerated and never incarcerated individuals^13^. To operationalize this, we assume that for people in prison, the ratio of their incarcerated to non-incarcerated contacts per unit time is approximately $\beta_{pp}:1$. This translates to approximately 1-5% of incarcerated individuals’ contacts being with non-incarcerated people. Additionally, because contact with staff and visitors is less frequent and generally of shorter duration than contact with other incarcerated individuals in the same cell or block/yard, we assume that the probability of infection upon contact with non-incarcerated individuals is approximately half of that upon contact with other incarcerated individuals. These assumptions result in a median $\beta_{pc}$ of 0.5, regardless of the value of $\beta_{pp}$, with an uncertainty distribution of Triangular(min=0.25, max=0.75, mode-0.5). We then impose symmetry as a constraint to obtain $\beta_{cp}$:

$${N_{p}\beta_{pc}= N}_{c}\beta_{cp}$$

$$\beta_{cp}=\beta_{pc}\frac{N_{p}}{N_{c}}$$

In a sensitivity analysis, we explore the effect of eliminating this mixing between incarcerated and non-incarcerated individuals (section 5).

*Model equations*

$$N_{p}=S_{p}+E_{p}+L_{p}+I_{p}+R_{p}$$

$$N_{r}=S_{r}+E_{r}+L_{r}+I_{r}+R_{r}$$

$$N_{d}=S_{d}+E_{d}+L_{d}+I_{d}+R_{d}$$

$$N_{n}=S_{n}+E_{n}+L_{n}+I_{n}+R_{n}$$

$$N=N_{p}+N_{r}+N_{d}+N_{n}$$

Prison

$$\frac{{dS}_{p}}{dt}=-{S_{p}(\beta}_{pp}\frac{I_{p}}{N_{p}}+\beta_{pc}\frac{I_{r}+I_{d}+I_{n}}{N_{r}+N_{d}+N_{n}})+q_{r}S_{r}+q_{d}S_{d}+q_{n}S_{n}-rS_{p}-{mu}_{p}S_{p}$$

$$\frac{{dE}_{p}}{dt}={(S}_{p}+\alpha L_{p}+\alpha R_{p})\left( \beta_{pp}\frac{I_{p}}{N_{p}}+\beta_{pc}\frac{I_{r}+I_{d}+I_{n}}{N_{r}+N_{d}+N_{n}} \right)-bE_{p}-\tau_{p}E_{p}+q_{r}E_{r}+q_{d}E_{d}+q_{n}E_{n}-rE_{p}-{mu}_{p}E_{p}$$

$$\frac{{dL}_{p}}{dt}=-\alpha L_{p}\left( \beta_{pp}\frac{I_{p}}{N_{p}}+\beta_{pc}\frac{I_{r}+I_{d}+I_{n}}{N_{r}+N_{d}+N_{n}} \right)+bE_{p}-{eL}_{p}+q_{r}L_{r}+q_{d}L_{d}+q_{n}L_{n}-rL_{p}-{mu}_{p}L_{p}$$

$$\frac{{dI}_{p}}{dt}=\tau_{p}E_{p}+{eL}_{p}-\delta_{p}I_{p}-\sigma I_{p}+\gamma R_{p}+q_{r}I_{r}+q_{d}I_{d}+q_{n}I_{n}-rI_{p}-{(mu}_{p}+{{mu}_{TB})I}_{p}$$

$$\frac{{dR}_{P}}{dt}=-\alpha R_{p}\left( \beta_{pp}\frac{I_{p}}{N_{p}}+\beta_{pc}\frac{I_{r}+I_{d}+I_{n}}{N_{r}+N_{d}+N_{n}} \right)+\delta_{p}I_{p}+\sigma I_{p}-\gamma R_{p}+q_{r}R_{r}+q_{d}R_{d}+q_{n}R_{n}-rR_{p}-{mu}_{p}R_{p}$$

Recent history of incarceration

$$\frac{{dS}_{r}}{dt}=-S_{r}\left( \beta_{cc}\frac{I_{r}+I_{d}+I_{n}}{N_{r}+N_{d}+N_{n}}+\beta_{cp}\frac{I_{p}}{N_{p}} \right)-q_{r}S_{r}+rS_{p}-\omega S_{r}-{mu}_{r}S_{r}$$

$$\frac{{dE}_{r}}{dt}={(S}_{r}+\alpha L_{r}+\alpha R_{r})\left( \beta_{cc}\frac{I_{r}+I_{d}+I_{n}}{N_{r}+N_{d}+N_{n}}+\beta_{cp}\frac{I_{p}}{N_{p}} \right)-bE_{r}-\tau_{r}E_{r}-q_{r}E_{r}+rE_{p}-\omega E_{r}-{mu}_{r}E_{r}$$

$$\frac{{dL}_{r}}{dt}=-\alpha L_{r}\left( \beta_{cc}\frac{I_{r}+I_{d}+I_{n}}{N_{r}+N_{d}+N_{n}}+\beta_{cp}\frac{I_{p}}{N_{p}} \right)+bE_{r}-{eL}_{r}-q_{r}L_{r}+rL_{p}-\omega L_{r}-{mu}_{r}L_{r}$$

$$\frac{{dI}_{r}}{dt}=\tau_{r}E_{r}+{eL}_{r}-\delta_{r}I_{r}-\sigma I_{r}+\gamma R_{r}-q_{r}I_{r}+rI_{p}-\omega I_{r}-{(mu}_{r}+{{mu}_{TB})I}_{r}$$

$$\frac{{dR}_{r}}{dt}=-\alpha R_{r}\left( \beta_{cc}\frac{I_{r}+I_{d}+I_{n}}{N_{r}+N_{d}+N_{n}}+\beta_{cp}\frac{I_{p}}{N_{p}} \right)+\delta_{r}I_{r}+\sigma I_{r}-\gamma R_{r}-q_{r}R_{r}+rR_{p}-\omega R_{r}-{mu}_{r}R_{r}$$

Distant history of incarceration

$$\frac{{dS}_{d}}{dt}=-S_{d}\left( \beta_{cc}\frac{I_{r}+I_{d}+I_{n}}{N_{r}+N_{d}+N_{n}}+\beta_{cp}\frac{I_{p}}{N_{p}} \right)-q_{d}S_{d}+\omega S_{r}-{mu}_{d}S_{d}$$

$$\frac{{dE}_{d}}{dt}={(S}_{d}+\alpha L_{d}+\alpha R_{d})\left( \beta_{cc}\frac{I_{r}+I_{d}+I_{n}}{N_{r}+N_{d}+N_{n}}+\beta_{cp}\frac{I_{p}}{N_{p}} \right)-bE_{d}-\tau_{d}E_{d}-q_{d}E_{d}+\omega E_{r}-{mu}_{d}E_{d}$$

$$\frac{{dL}_{d}}{dt}=-\alpha L_{d}\left( \beta_{cc}\frac{I_{r}+I_{d}+I_{n}}{N_{r}+N_{d}+N_{n}}+\beta_{cp}\frac{I_{p}}{N_{p}} \right)+bE_{d}-{eL}_{d}-q_{d}L_{d}+\omega L_{r}-{mu}_{d}L_{d}$$

$$\frac{{dI}_{d}}{dt}=\tau_{d}E_{d}+{eL}_{d}-\delta_{d}I_{d}-\sigma I_{d}+\gamma R_{d}-q_{d}I_{d}+\omega I_{r}-{(mu}_{d}+{{mu}_{TB})I}_{d}$$

$$\frac{{dR}_{d}}{dt}=-\alpha R_{d}\left( \beta_{cc}\frac{I_{r}+I_{d}+I_{n}}{N_{r}+N_{d}+N_{n}}+\beta_{cp}\frac{I_{p}}{N_{p}} \right)+\delta_{d}I_{d}+\sigma I_{d}-\gamma R_{d}-q_{d}R_{d}+\omega R_{r}-{mu}_{d}R_{d}$$

Never incarcerated

$$\frac{{dS}_{n}}{dt}=vi\left( 1-\left( 1-e^{-(\beta_{cc}\frac{I_{r}+I_{d}+I_{n}}{N_{r}+N_{d}+N_{n}} + \beta_{cp}\frac{I_{p}}{N_{p}})\left( 15 \right)} \right) \right)\left( {{mu}_{p}N_{p}+{mu}_{r}N_{r}+{mu}_{d}N_{d}+mu}_{n}N_{n}+{mu}_{TB}\left( I_{p}+I_{r}+I_{d}+I_{n} \right) \right)-S_{n}\left( \beta_{cc}\frac{I_{r}+I_{d}+I_{n}}{N_{r}+N_{d}+N_{n}}+\beta_{cp}\frac{I_{p}}{N_{p}} \right)-q_{n}S_{n}-{mu}_{n}S_{n}$$

$$\frac{{dE}_{n}}{dt}=\left. vi*{prop}_{E}\left( 1-e^{-(\beta_{cc}\frac{I_{r}+I_{d}+I_{n}}{N_{r}+N_{d}+N_{n}} + \beta_{cp}\frac{I_{p}}{N_{p}})\left( 15 \right)} \right) \right.\left( {{mu}_{p}N_{p}+{mu}_{r}N_{r}+{mu}_{d}N_{d}+mu}_{n}N_{n}+{mu}_{TB}\left( I_{p}+I_{r}+I_{d}+I_{n} \right) \right)+{(S}_{n}+\alpha L_{n}+\alpha R_{n})\left( \beta_{cc}\frac{I_{r}+I_{d}+I_{n}}{N_{r}+N_{d}+N_{n}}+\beta_{cp}\frac{I_{p}}{N_{p}} \right)-bE_{n}-\tau_{n}E_{n}-q_{n}E_{n}-{mu}_{n}E_{n}$$

$$\frac{{dL}_{n}}{dt}=\left. vi*{(1-prop}_{E})\left( 1-e^{-(\beta_{cc}\frac{I_{r}+I_{d}+I_{n}}{N_{r}+N_{d}+N_{n}} + \beta_{cp}\frac{I_{p}}{N_{p}})\left( 15 \right)} \right) \right.\left( {{mu}_{p}N_{p}+{mu}_{r}N_{r}+{mu}_{d}N_{d}+mu}_{n}N_{n}+{mu}_{TB}\left( I_{p}+I_{r}+I_{d}+I_{n} \right) \right)-\alpha L_{n}\left( \beta_{cc}\frac{I_{r}+I_{d}+I_{n}}{N_{r}+N_{d}+N_{n}}+\beta_{cp}\frac{I_{p}}{N_{p}} \right)+bE_{n}-{eL}_{n}-q_{n}L_{n}-{mu}_{n}L_{n}$$

$$\frac{{dI}_{n}}{dt}=\tau_{n}E_{n}+{eL}_{n}-\delta_{n}I_{n}-\sigma I_{n}+\gamma R_{n}-q_{n}I_{n}-{(mu}_{n}+{{mu}_{TB})I}_{n}$$

$$\frac{{dR}_{n}}{dt}=-\alpha R_{n}\left( \beta_{cc}\frac{I_{r}+I_{d}+I_{n}}{N_{r}+N_{d}+N_{n}}+\beta_{cp}\frac{I_{p}}{N_{p}} \right)+\delta_{n}I_{n}+\sigma I_{n}-\gamma R_{n}-q_{n}R_{n}-{mu}_{n}R_{n}$$

2. **Parameter calibration and uncertainty analysis**

Calibration targets are described in **Tables S1-S2**. We generated joint uncertainty distributions for calibration targets and non-calibrated parameters as described in **Table S4**, using Latin Hypercube Sampling. For tuberculosis incidence targets, we generated uncertainty distributions as follows. We retrieved population-wide notifications data and incidence estimates with uncertainty intervals from the 2023 WHO Global Tuberculosis Report. WHO incidence estimates were derived using notification data adjusted by a standard factor that varied by year and country; in 2020-2022 incidence was estimated using country- or region-specific dynamic models^14^. As we did not have the raw posterior distributions for incidence estimates, we assumed the underlying data (and the ratio of incidence to notifications) followed a normal distribution, with the main estimate and upper bound representing the mean and 97.5 percentile, respectively, for each country and year. We then sampled quantiles to apply to the distribution across all years to generate uncertainty in the calibration target of population TB incidence.

We obtained uncertainty distributions for prison incidence targets as follows. For Brazil and Colombia, we sourced posterior distributions directly from a recent study integrating notifications data with empirical incidence and prevalence estimates from primary active case finding studies^9^. For the remaining countries where no such empirical studies have been conducted (Peru, Mexico, El Salvador, and Argentina), we estimated incidence from notifications data by calculating and applying a regional case detection ratio (CDR) from countries with primary active finding studies (Brazil, Colombia, and Chile). We sampled uniformly from the posterior CDR distribution shown in **Figure S2**. For all countries, we fit only to prison incidence estimates for years when prison notifications data were available. We induced an approximate correlation of 0.3 between the uncertainty distributions for population-wide and prison incidence.

For each of 3000 sampled sets of calibration targets and non-calibrated parameters, we ran optimization algorithms to fit the remaining parameters in a two-step process. **Table S3** shows which parameters were calibrated in the first step versus second step. First, we calibrated incarceration-related parameters using a simple population transition model that distinguished those in prison by prior incarceration history (**Figure S3**). This enabled us to calibrate different prison entry rates based on incarceration history to fit to recidivism data targets. We assumed the same release rate for all individuals in prison regardless of incarceration history. We then fixed incarceration-related parameters for the second step of calibration, using the main meta-population dynamic model to calibrate tuberculosis-related parameters.

We conducted calibration with *optim* in R using limited memory BFGS, a quasi-Newton algorithm, with box constraints. The first step of calibration minimized the mean squared percentage error across incarceration-related data targets. The loss function gave extra weight to the initial and final prevalence of incarceration given the importance of a good fit to these for the historical counterfactual analysis and future projections. The loss function was the mean of the following five errors:

1) Mean squared percentage error of incarceration prevalence $P$ at all time points:

$$\frac{1}{n}\sum_{t=1}^{n} \left( \frac{\hat{P_{t}}-P_{t}}{P_{t}} \right)^{2}$$

2) Squared percentage error of incarceration prevalence at baseline $P_{0}$:

$$\left( \frac{\hat{P_{0}}-P_{0}}{P_{0}} \right)^{2}$$

3) Mean squared percentage error of incarceration prevalence in 2019 and at the final time point:

$$\frac{1}{2}\left( \left( \frac{\hat{P_{2019}}-P_{2019}}{P_{2019}} \right)^{2}+\left( \frac{\hat{P_{n}}-P_{n}}{P_{n}} \right)^{2} \right)$$

4) Mean squared percentage error of recidivism $R$ at all time points:

$$\frac{1}{n}\sum_{t=1}^{n} \left( \frac{\hat{R_{t}}-R_{t}}{R_{t}} \right)^{2}$$

5) Mean squared percentage error of prison admissions $A$ at all time points:

$$\frac{1}{n}\sum_{t=1}^{n} \left( \frac{\hat{A_{t}}-A_{t}}{A_{t}} \right)^{2}$$

For final incarceration prevalence (error #3), we took the average of the mean squared percentage error in 2019 and the mean squared percentage error in the most recent year with data (2022 or 2023 depending on the country).

The second step of calibration minimized the mean percentage error across tuberculosis-related data targets. The loss function for this second step was the mean of the following four errors:

1) Mean absolute percentage error of TB incidence rate in prison, $Ip$, at all time points:

$$\frac{1}{n}\sum_{t=1}^{n} \left| \frac{\hat{{Ip}_{t}}-{Ip}_{t}}{{Ip}_{t}} \right|$$

2) Mean absolute percentage error of TB incidence rate across all strata combined, $Ic$, at all time points:

$$\frac{1}{n}\sum_{t=1}^{n} \left| \frac{\hat{{Ic}_{t}}-{Ic}_{t}}{{Ic}_{t}} \right|$$

3) Mean absolute percentage error of TB notifications rate in prison, $Dp$, at all time points:

$$\frac{1}{n}\sum_{t=1}^{n} \left| \frac{\hat{{Dp}_{t}}-{Dp}_{t}}{{Dp}_{t}} \right|$$

4) Mean absolute percentage error of TB notifications rate across all strata combined, $Dc$, at all time points:

$$\frac{1}{n}\sum_{t=1}^{n} \left| \frac{\hat{{Dc}_{t}}-{Dc}_{t}}{{Dc}_{t}} \right|$$

For many parameters, final calibrated values were highly correlated with starting values; furthermore, different combinations of parameter values generated similar fits to the data, indicating model non-identifiability. Against this backdrop, we sought to maximize representation of the possible parameter space by sampling from joint uncertainty distributions for starting values of calibrated parameters, generated through Latin Hypercube Sampling.

Country-specific priors and posterior distributions for calibrated parameters are shown in **Figure S4.** Time-varying parameters are shown in **Figure S5.** Model fits to calibration targets are presented in **Figures S6-S7**.

3. **Model validation**

After model calibration, we performed model validation with additional data that were not used as calibration targets, described below. For each data source, we specify their degree of independence from model construction (Eddy et al. 2012).

| Data | Availability | Source | Data Type | Independence |
| --- | --- | --- | --- | --- |
| Tuberculosis deaths | All countries, 2000-2019 (through 2022 in Brazil) | WHO | Estimated using administrative data | Mostly independent: estimates were based on vital registration cause-of-death data. For some countries, these data were used to estimate tuberculosis incidence (a calibration target) in the years 2020-2022. |
| Prevalence of tuberculosis in prisons | Brazil and Colombia, varying years | Nogueira et al. 2012; Carbone et al. 2015; Lemos et al. 2009; Estevan et al. 2013; Abrahao et al. 2006; Alarcón-Robayo et al. 2016; Guerra et al. 2019 | Primary data from empirical studies | Partially dependent: most of these data were used in the Bayesian hierarchical model in Martinez & Warren et al. 2024 to generate estimates of tuberculosis incidence in prisons (a calibration target). |
| Prevalence of latent tuberculosis infection in prisons | Brazil and Colombia, varying years | Nogueira et al. 2012; Carbone et al. 2015; Lemos et al. 2009; Estevan et al. 2013; Abrahao et al. 2006; Rueda et al. 2014; Guerra et al. 2019 | Primary data from empirical studies | Independent: data were not used for building any part of the model. |

Consistency between model outputs and validation data is shown in **Figure S8** and described below.

Tuberculosis deaths. The estimates and confidence intervals for the tuberculosis mortality rate fall within the uncertainty bounds of our model outputs and generally exhibit similar trends over time for all countries, with some inconsistencies in Mexico and El Salvador. In Mexico, our model underestimates tuberculosis deaths from 2000-2003, which likely reflects our simplifying assumption of a constant tuberculosis mortality rate over time. In El Salvador, our model predicts an increase in tuberculosis mortality after 2010, a trend that was not observed in the data, despite increasing tuberculosis incidence during that period. This is likely due to our model’s lack of age structure, as the increase in incidence was driven largely by increasing cases in prisons, largely among men under 45 who are less likely to die from tuberculosis. It is also possible, however, that some tuberculosis deaths in prisons were not captured due to underreporting and poor quality of cause-of-death information for deaths in prisons. Nevertheless, due to the relatively low rates of tuberculosis mortality, these inconsistencies are unlikely to substantially impact our model results and conclusions. Moreover, we do not report estimates of tuberculosis mortality in our manuscript findings.

Prevalence of tuberculosis in prisons. In Brazil and Colombia, our model estimates of tuberculosis prevalence in prisons are generally concordant with empirical studies conducted in different prisons and regions. In Brazil, the highest estimate of tuberculosis prevalence came from a study conducted in a prison hospital (Lemos et al. 2009), where the population may have higher prevalence of tuberculosis risk factors. We were unable to find empirical studies on tuberculosis prevalence in prisons in the other four countries.

Prevalence of latent tuberculosis infection (LTBI) in prisons. Our model estimates of the prevalence of LTBI in prisons are largely consistent with empirical studies conducted in different prisons and regions in Brazil. The lowest estimate of LTBI prevalence in prisons came from a study in Mato Grosso do Sul (Carbone et al. 2015), where the authors commented that the difference observed compared to previous studies was likely due to lower rates of tuberculosis in the state. In Colombia, two studies found higher prevalence of LTBI in prisons than estimated by our model. In both studies, the study population had a higher prevalence of incarceration history than the national prison population (23.6% and 30.4%, compared to 17% nationally). Therefore, greater exposure to incarceration may have contributed to the higher LTBI prevalence observed compared to our model estimates.

Due to the dearth of empirical data on tuberculosis and incarceration in Latin America, we were limited in our ability to perform external model validation with independent data sources from all six countries. Our model should be assessed for consistency with external data from future empirical studies and other independent sources as they become available.

4. **Historical counterfactual scenarios**

*Transmission population attributable fraction (tPAF)*

To estimate the tPAF in 2019, we simulated a scenario where, between 1990 and 2009, prison entry rates gradually decreased to zero and release rates gradually increased, such that incarceration prevalence was approximately zero by 2009, with no new exposure to incarceration occurring thereafter. We then calculated the PAF for incident cases in the year 2019, ten years later to account for lagged effects from prior exposure to incarceration and onward transmission.

5. **Sensitivity analyses and meta-modeling**

*Latent infection upon birth into model*

In the main analysis, we assume a 1:6 ratio of early to late latent infection among individuals born into the model at age 15 with latent infection, based on evidence of a median incubation period for *Mycobacterium tuberculosis* infection of under two years^15^. In a sensitivity analysis, we test a 1:2 ratio of early to late latent infection, based on prior work suggesting an incubation period up to five years^16^.

*Progression and diagnosis rates in recent incarceration history stratum*

In the main analysis, we assume that individuals with recent incarceration history have higher disease progression rates and lower diagnosis rates than individuals with distant or no incarceration history. We remove these differences in a sensitivity analysis.

*Effective contact rates in prison over time*

In the main analysis, we assume that the effective contact rate in prison can change over time for some countries, based on observed changes over time in prison tuberculosis rates that could not be accounted for solely by changes in case detection or incarceration dynamics. We performed a sensitivity analysis holding the effective contact rate in prison constant over time.

*Assortative mixing*

In our main analysis, we assume proportionate mixing in the community, with $\beta_{cc}$ representing the effective contact rate for all intra- and inter-stratum mixing in the community (i.e. among those never incarcerated, those with recent history of incarceration, and those with distant history of incarceration). In a sensitivity analysis, we implement assortative mixing in the community by incarceration history. We consider those with recent or distant history of incarceration as one group (formerly incarcerated or “f”), with the other group being never incarcerated (“n”). This results in the following contact matrix:

$$\boldsymbol{\beta}=\left[ \begin{matrix} \begin{matrix} \beta_{ff} & \beta_{fn} \end{matrix} \\ \begin{matrix} \beta_{nf} & \beta_{nn} \end{matrix} \end{matrix} \right]$$

These are operationalized in the model equations as follows (examples for Susceptibles shown):

$$\frac{{dS}_{r}}{dt}=-S_{r}\left( \beta_{ff}\frac{I_{r}+I_{d}}{N_{r}+N_{d}}+\beta_{fn}\frac{I_{n}}{N_{n}}+\beta_{cp}\frac{I_{p}}{N_{p}} \right)\ldots$$

$$\frac{{dS}_{d}}{dt}=-S_{d}\left( \beta_{ff}\frac{I_{r}+I_{d}}{N_{r}+N_{d}}+\beta_{fn}\frac{I_{n}}{N_{n}}+\beta_{cp}\frac{I_{p}}{N_{p}} \right)\ldots$$

$$\frac{{dS}_{n}}{dt}=-S_{n}\left( \beta_{nn}\frac{I_{n}}{N_{n}}{+\beta}_{nf}\frac{I_{r}+I_{d}}{N_{r}+N_{d}}+\beta_{cp}\frac{I_{p}}{N_{p}} \right)\ldots$$

Under proportionate mixing, contacts with other community members are distributed based on the proportion of the population in each group, where $N_{i}$ is the number of individuals in group I, such that the contact matrix is as follows:

$$\boldsymbol{\beta}_{\boldsymbol{proportionate}}=\left[ \begin{matrix} \begin{matrix} {\frac{N_{f}}{{N_{f}+N}_{n}}\beta}_{cc} & {\frac{N_{n}}{{N_{f}+N}_{n}}\beta}_{cc} \end{matrix} \\ \begin{matrix} {\frac{N_{f}}{{N_{f}+N}_{n}}\beta}_{cc} & {\frac{N_{n}}{{N_{f}+N}_{n}}\beta}_{cc} \end{matrix} \end{matrix} \right]$$

To impose assortative mixing, we assume $\beta_{ff\_assortative}=m\beta_{ff\_proportionate}$. We set a constraint to maintain the same total contact rate for each group as under the proportionate mixing assumption, such that $\beta_{ff\_assortative}+\beta_{fn\_assortative}=\beta_{nn\_assortative}+\beta_{nf\_assortative}=\beta_{cc}$. We note that this results in the following limits for $m$:

$$1< m< \frac{N_{f}+N_{n}}{N_{f}}$$

We assume symmetry for between-group interactions, such that ${N_{f}\beta}_{fn}={N_{n}\beta}_{nf}.$ We can then obtain the complete contact matrix as follows:

$$\beta_{ff}={\frac{N_{f}m}{{N_{f}+N}_{n}}\beta}_{cc}$$

$$\beta_{fn}=\beta_{cc}-\beta_{ff}$$

$$\beta_{nf}={\frac{N_{f}}{N_{n}}\beta}_{fn}$$

$$\beta_{nn}=\beta_{cc}-\beta_{nf}$$

For this sensitivity analysis, we set $m=3$, such that within-group mixing among formerly incarcerated individuals is three times the amount relative to what would be expected under proportionate mixing.

*Direct prison-community mixing*

In the main analysis, we allow low levels of mixing between incarcerated and non-incarcerated individuals to represent interactions with prison staff and visitors. We deactivate this mixing in a sensitivity analysis.

For all sensitivity analyses, we re-assess model fit and re-calibrate if the error increases by more than 10%. Results for all sensitivity analyses are shown in **Figure S9.**

*Meta-modeling*

We performed linear regression meta-modeling to infer which parameters, calibrated or non-calibrated, were most strongly associated with variation in our excess burden estimates. We included results from all countries’ fitted parameter sets to obtain generalizable inferences. Given this data structure, we used a multi-level model in order to account for country-level effects. We first harmonized the parameters to enable more intuitive interpretation. Since time-varying parameters changed over different intervals across countries, we transformed parameters governing changes over time into parameters representing net percentage change from baseline. We then scaled all parameters so that meta-model coefficients would represent the change in outcome associated with one standard deviation change in a given parameter^17^. We excluded parameters that were not specified for all countries, such as the relative diagnosis rate in prisons vs. outside at baseline, the change in the prison diagnosis rate, or the change in the prison effective contact rate. We included country-level varying intercepts and varying slopes for a subset of parameters, chosen using the model Akaike Information Criterion (AIC). We also included tuberculosis notification rates in prisons and in the general population as country-level covariates. Our model had a marginal R^2^ of 0.90 and a conditional R^2^ of 0.98.

Meta-modeling showed that most of the variation in excess burden estimates can be attributed to country-level differences **(Table S10)**. Additional variation across fitted parameter sets for each country can be largely explained by parameters governing tuberculosis incidence in and out of prison and parameters underlying incarceration growth and dynamics. Specifically, parameter values leading to greater disparity in tuberculosis incidence in prisons versus outside, greater increase in incarceration prevalence, or higher prison turnover rates, were associated with higher excess burden estimates.

6. **Future projections**

For the “continue trends” scenario, implemented for all countries except El Salvador (see below), we calculate the net percentage change in entry and release rates between 2013 and 2023 and simulate a continuation of that change between 2024 and 2034. Here are the median net percentage changes in entry and release rates between 2013 and 2023 for each country:

|  | Entry rates *q_n_* and *q_d_* (%) | Release rate *r* (%) |
| --- | --- | --- |
| Argentina | 37 | 0 |
| Brazil | 0 | 4 |
| Colombia | -19 | 0 |
| Mexico | -40 | -27 |
| Peru | -8 | -27 |

We note that while incarceration prevalence exhibited a net decline in Mexico between 2013 and 2023, the decrease in the release rate led to an increase in within-prison incidence. This may explain why projected population tuberculosis incidence in 2034 for Mexico is higher in the “continue trends” scenario compared to the “stable” scenario of constant entry and release rates. Moreover, although incarceration prevalence underwent a net increase in Brazil between 2013 and 2023, entry and release rates themselves exhibited a reversal in direction of change during the same period, leading to net changes in entry and release rates close to zero.

For decarceration scenarios, reductions in average duration of incarceration were operationalized by increasing the release rate, given their reciprocal relationship. To achieve 25% or 50% reductions in duration, the release rate was gradually increased by 33% (1/0.75 - 1) or 100% (1/0.5 – 1), respectively, between 2024 and 2034. Incarceration prevalence in 2034 under each scenario is shown in **Table S13.**

*El Salvador*

Since March 2022, El Salvador has maintained a state of emergency implemented as part of a “war on gangs”. As of January 2024, over 75,000 people have been arrested, and the latest official figures from August 2023 indicate that less than 10% of those detained have been released^18,21^. As of February 2024, the state of emergency continues to be renewed^18,19^.

We operationalized the state of emergency in our model as follows. Based on updates from the Salvadoran Legislative Assembly^18^, we assume that the rate of arrests peaked in the first three months of the state of emergency (March-June 2022) and subsequently exhibited exponential decay until August 2023, when it stabilized at a level higher than before the state of emergency. Based on statements by government officials^20,21^, we assume that the release rate dropped at the start of the state of emergency and has remained constant since. Details on parameterization are provided in **Table S14**.

For future projections, we simulate the following scenarios between 2024 and 2034:

1. Continue state of emergency: entry and release rates remain at their current estimated level (as of January 2024) for ten years.
2. Gradual passive abatement: entry and release rates gradually return to pre-state-of-emergency levels by 2034.
3. Active 10-year reversion: incarceration prevalence reverts to its approximate pre-emergency level by 2034. This is achieved through entry rates returning to pre-emergency values, and the release rate increasing to 1.25 times its pre-emergency value, by 2025. Entry and release rates are then held constant for the rest of the period.
4. Active 5-year reversion: incarceration prevalence reverts to its approximate pre-emergency level by 2029, with continued decarceration thereafter. This is achieved through entry rates returning to pre-emergency values, and the release rate increasing to twice its pre-emergency value, by 2025. Entry and release rates are then held constant for the rest of the period.
5. Active 2-year reversion: incarceration prevalence reverts to its approximate pre-emergency level by 2026, with continued decarceration thereafter. This is achieved through entry rates returning to 1990 values, and release rates increasing to 4.75 times their pre-emergency value, by 2025. Entry rates are maintained for the rest of the period. In 2026 the release rate returns to its 1990 value and is constant for the rest of the period.

Incarceration prevalence in 2034 under each scenario is shown in **Table S15.**

**Table S1. Data availability and sources for incarceration-related calibration targets.**

|  | Argentina | Brazil | Colombia | El Salvador | Mexico | Peru |
| --- | --- | --- | --- | --- | --- | --- |
| Prevalence of incarceration* | 1992, 1995, 1997-2022 | 1990-2023 | 1990-2022 | 1990, 1995, 2000-2023 | 1990, 1992, 1993, 1995, 1998-2008^#^, 2010-2023 | 1990, 1995, 1997-2023 |
| Recidivism** | 2002-2021 | 2013 | 2015-2021 | 2013, 2017-2019 | 2017-2019 | 2016-2023 |
| Prison admissions*** | 2004-2022 | 2016-2022 | 2009-2021 | 1990-2020 | 2010-2022 | 2000, 2005, 2010-2021 |
| Source | El Sistema Nacional de Estadísticas sobre Ejecución de la Pena (SNEEP)^22^; Ministry of Justice information request | Sistema de Informações do Departamento Penitenciário Nacional (SISDEPEN)^23^; Sanguinetti et al. 2014^24^; Bergman et al. 2015^25^; de Azevedo RD and Cifali AC 2017^26^ | Instituto Nacional Penitenciario y Carcelario (INPEC)^27^; Ministry of Justice information request | Dirección General de Centros Penales (DGCP), retrieved Jan. 2020^28^; Instituto Universitario de Opinión Pública (IUDOP)^29^; Bergman et al. 2015^25^; La Prensa Gráfica 2019^30^ | Instituto Nacional de Estadística y Geografia (INEGI)^31^ | El Instituto Nacional Penitenciario (INPE)^32^; International Committee of the Red Cross (ICRC)^33^ |

^#^every other year

*For Colombia and Mexico, where incarceration prevalence was higher in 1990 than in years before and after, we fit a smooth spline to the data and sampled from a uniform distribution ranging between the observed prevalence and the spline estimate to get the calibration target for 1990 prevalence. Resulting model fits to incarceration prevalence are shown in **Figure S6**.

**For all countries except Mexico, data on recidivism were reported as the cross-sectional percent of the prison population with prior incarceration history. In Mexico, data on recidivism were reported as the percent of all prison admissions where the individual was previously incarcerated.

***Admissions data in Argentina from 2004-2019 excluded those who were detained and released in the same year. We used admissions data from 2020-2022, which included this subset of people, to estimate total admissions for 2004-2019, assuming a constant ratio of total admissions to the number admitted and released within the same year. Total admissions in Brazil were estimated using data from facilities holding >80% of the prison population in each semester, assuming missingness-at-random.

**Table S2. Data availability and sources for tuberculosis-related calibration targets.**

|  | Argentina | Brazil | Colombia | El Salvador | Mexico | Peru | Source |
| --- | --- | --- | --- | --- | --- | --- | --- |
| Total TB notification rate | 1990-2022 | 1990-2022 | 1990-2022 | 1990-2022 | 1990-2022 | 1990-2022 | WHO Global TB Report |
| Total TB incidence rate | 2000-2022 | 2000-2022 | 2000-2022 | 2000-2022 | 2000-2022 | 2000-2022 | WHO Global TB Report |
| Prison TB notification rate | 2014-2022 | 2007-2022 | 2013-2022 | 2002-2022 | 2012-2022 | 2010-2022 | Country-level case notifications collated by PAHO |
| Prison TB incidence rate* | 2014-2022 | 2007-2019 | 2007-2019 | 2002-2022 | 2012-2022 | 2010-2022 | Martinez & Warren et al. 2023 |

*We directly used posterior distributions of prison incidence for Brazil and Colombia, where empirical active case-finding studies have been conducted. For the other four countries, we used a regional case detection ratio (CDR) to generate prison incidence estimates for calibration. The posterior CDR distribution, from which we sampled randomly, is shown in **Figure S2**.

**Table S3. Description of model parameters.** Some parameters indicated as time-varying are only time-varying in select countries (see Table S5). Parameters without subscripts are assumed to be equal across strata. Triangular distributions are specified as Tri(minimum, maximum, mode).

| Parameter | Description | Calibrated, sampled, or fixed? | Time-varying? | Prior | Source | If calibrated, first or second step |
| --- | --- | --- | --- | --- | --- | --- |
| $q_{r}$ | Entry rate into prison from **r** stratum | Calibrated | Yes | Country-specific | Calibrated | First step |
| $q_{d} , q_{n}$ | Entry rate into prison from **d** and **n** strata | Calibrated as ratio relative to $q_{r}$ | Yes | Country-specific | Calibrated | First step |
| $r$ | Release rate from prison | Calibrated | Yes | Country-specific | Calibrated | First step |
| $\omega$ | Rate of transition from **r** stratum to **d** stratum | Fixed | No | 1/7 | ^3,11^ |  |
| $\beta_{pp}$ | Effective contact rate for within-prison transmission | Calibrated | Yes | Country-specific | Calibrated | Second step |
| $\beta_{cc}$ | Effective contact rate for transmission among individuals outside prison | Calibrated as ratio relative to $\beta_{pp}$ | Yes | Country-specific | Calibrated | Second step |
| $\beta_{pc} , \beta_{cp}$ | Effective contact rate for transmission between individuals in prison and outside prison; calculated as $\beta_{cc}$at baseline * prevalence of incarceration * uncertainty factor | Sampled (uncertainty factor) | No | Uncertainty factor ~ Tri(0.5, 1.5, 1) | Assumed |  |
| $\tau_{p}$ | Fast progression rate from early latent to infectious in prison | Calibrated | No | Country-specific | Calibrated | Second step |
| $\tau_{d} ,\tau_{n}$ | Fast progression rate from early latent to infectious in **d** and **n** strata | Calibrated as ratio relative to $\tau_{p}$ | No | Country-specific | Calibrated | Second step |
| $\tau_{r}$ | Fast progression rate from early latent to infectious in **r** stratum; calculated as weighted average of $\tau_{p}$ and $\tau_{d} ,\tau_{n}$ | Sampled (weight) | No | Weight ~ Unif(0, 1) | Assumed |  |
| $\delta_{d} , \delta_{n}$ | Diagnosis rate in **d** and **n** strata | Calibrated | Yes | Country-specific | Calibrated | Second step |
| $\delta_{p}$ | Diagnosis rate in prison | Calibrated as ratio relative to $\delta_{d} , \delta_{n}$; equal to $\delta_{d} , \delta_{n}$ at baseline in Peru, Mexico, & Brazil | Yes | Country-specific | Calibrated | Second step |
| $\delta_{r}$ | Diagnosis rate in **r** stratum; calculated as weighted average of $\delta_{p}$ and $\delta_{d} , \delta_{n}$ | Sampled (weight) | Yes | Weight ~ Unif(0, 1) | Assumed |  |
| $b$ | Transition rate from early to late latent | Sampled | No | Tri(0.75, 1, 0.875) | ^34^ |  |
| $e$ | Slow progression rate from late latent to infectious | Fixed | No | 0.000594 | ^34^ |  |
| $\alpha$ | Relative risk of reinfection for late latent and recovered individuals | Sampled | No | Lognormal(-1.56, 0.03) | ^35^ |  |
| $\gamma$ | Rate of relapse from R to I | Fixed | No | 0.01 | ^36^ |  |
| ${mu}_{TB}$ | TB mortality rate, assuming 10-50% prevalence of smear-positive TB | Sampled | No | Unif(0.061, 0.21) | ^37^ |  |
| $\sigma$ | Self-cure rate | Sampled | No | Unif(0.14, 0.18) | ^37^ |  |
| ${mu}_{d} , {mu}_{n}$ | Mortality rate in **d** and **n** strata | Fixed | Yes | Country-specific | ^38^ |  |
| ${mu}_{p}$ | Mortality rate in prison | Sampled as ratio relative to ${mu}_{d} , {mu}_{n}$ | No (constant ratio) | Rate ratio ~ $N$(0.65, 0.11) | ^3^ |  |
| ${mu}_{r}$ | Mortality rate in **r** stratum | Sampled as ratio relative to ${mu}_{d} , {mu}_{n}$ | No (constant ratio) | Rate ratio ~ $N$(1.05, 0.18) | ^3^ |  |
| $vi$ | Ratio of birth rate to death rate | Fixed | Yes | Country-specific | ^38^ |  |
| ${prop}_{E}$ | Proportion of people with latent TB born into the model at age 15 with early latent infection (E) | Fixed | No | 1/7 | ^15^ |  |

**Table S4. Uncertainty distributions for calibration targets.** CDR, case detection ratio; UI, uncertainty interval. Triangular distributions are specified as Tri(minimum, maximum, mode).

|  | Argentina | Brazil | Colombia | El Salvador | Mexico | Peru |
| --- | --- | --- | --- | --- | --- | --- |
| Total incidence^#^ | Notifications * $N$(1.16, 0.093) | Notifications * $N$(1.16, 0.095) | Notifications * $N$(1.27, 0.16) | Notifications * $N$(1.25, 0.17) | Notifications * $N$(1.26, 0.16) | Notifications * $N$(1.29, 0.21) |
| Prison incidence | Prison notifications / regional CDR: 0.54 (95% UI, 0.20-0.96) | Posterior distribution from Martinez, Warren, et al. 2023 | Posterior distribution from Martinez, Warren, et al. 2023 | Prison notifications / regional CDR: 0.54 (95% UI, 0.20-0.96) | Prison notifications / regional CDR: 0.54 (95% UI, 0.20-0.96) | Prison notifications / regional CDR: 0.54 (95% UI, 0.20-0.96) |
| Recidivism^+^ | Recidivism odds * Tri(0.5, 1.5, 1.0) | | | | | |
| Admissions | Admissions rate * Tri(0.75, 1.25, 1) | | | | | |

^#^Incidence factor calculated by dividing WHO incidence estimates by notifications; summary statistics are across all years but factors used in analysis were country- and year-specific

+Uncertainty distribution generated using odds to avoid exceeding 1.0

**Table S5. Intervals and direction of change in time-varying parameters.** Decisions were informed by observed tuberculosis data and prison admission/release data. Excludes changes due to COVID-19 pandemic (see Table S6) or El Salvador’s state of emergency (see Table S14).

|  | Argentina | Brazil | Colombia | El Salvador | Mexico | Peru |
| --- | --- | --- | --- | --- | --- | --- |
| Prison entry rates $q_{r} , q_{d} , q_{n}$ | Increases [1992-2000), [2000-2002), [2011-2016) | Increases [1990, 2020); decreases [2021, 2023) | Increases [1992, 2009); decreases [2009, 2015) | Increases [2005, 2007) and [2013, 2015); decreases [2018, 2020) | Increases [1995, 2002); decreases [2014, 2016); increases [2019, 2021) | Increases [1995, 1999); decreases [1999, 2003) and [2011, 2014) |
| Prison release rate $r$ | Increases [2004, 2006) | Decreases [1990, 2015); increases [2015, 2020) | Decreases [1992, 2009) and [2009, 2012) | Decreases [1995, 2002) | Decreases [1995, 2002) and [2013, 2017) | Decreases [2001, 2007) and [2009, 2014) |
| Effective contact rate in community $\beta_{cc}$ | Decreases [2002, 2010); increases [2013, 2020) | Decreases [1990, 2015) | Decreases [1990, 2020) | Decreases [1990, 2000) | Increases [1994, 1996); decreases [1996, 2002); increases [2006, 2020) | Decreases [1990, 2000) and [2000, 2020) |
| Effective contact rate in prison $\beta_{pp}$ | Increases [2015, 2020) | Increases [2015, 2018) | No change | Increases [2005, 2017); decreases [2018, 2022.25) | Decreases [2016, 2020) | No change |
| Diagnosis rates in community $\delta_{r} ,\delta_{d} , \delta_{n}$ | Increases [1992, 2000) | Increases [1990, 2015) | Increases [1990, 2020) | Increases [1990, 2000) | Increases [1995, 1998) | Increases [1990, 2020) |
| Diagnosis rate in prison $\delta_{p}$ | No change | No change | No change | Increases [2012, 2017); elevated [2017, 2020.25)* | Increases [2012, 2020) | No change |

*operationalized as a step function where $\delta_{p}$ is 1.3-2.5 times higher; informed by El Salvador’s National Strategic Plan for Tuberculosis Control, 2017-2021.

**Table S6. Changes in model parameters during COVID-19 pandemic.** All changes were implemented as step functions, with parameters increasing or decreasing by a calibrated or sampled factor during the period indicated.

| Country | Period | Parameter | Parameter description | Change | Prior if sampled, or calibrated? |
| --- | --- | --- | --- | --- | --- |
| Argentina | [2020.25, 2022) | $q_{r} , q_{d} , q_{n}$ | Prison entry rates | Decrease | Calibrated |
|  |  | $r$ | Prison release rate | Increase | Inverse of above |
|  |  | $\beta_{cp}{,\beta}_{pc}, \beta_{cc}$ | Effective contact rates in community | Decrease | Tri(0.6, 1.0, 0.8) |
|  | [2020.25, 2021.5) | $\delta_{p} , \delta_{r} ,\delta_{d} , \delta_{n}$ | Diagnosis rates | Decrease | Calibrated |
| Brazil | [2020.25, 2021) | $q_{r} , q_{d} , q_{n}$ | Prison entry rates | Decrease | Calibrated |
|  |  | $r$ | Prison release rate | Increase | Inverse of above |
|  |  | $\beta_{cp}{,\beta}_{pc}, \beta_{cc}$ | Effective contact rates in community | Decrease | Tri(0.6, 1.0, 0.8) |
|  | [2020.25, 2021.5) | $\delta_{p} , \delta_{r} ,\delta_{d} , \delta_{n}$ | Diagnosis rates | Decrease | Calibrated |
| Colombia | [2020.25, 2022) | $q_{r} , q_{d} , q_{n}$ | Prison entry rates | Decrease | Calibrated |
|  | [2020.25, 2021) | $\beta_{cp}{,\beta}_{pc}, \beta_{cc}$ | Effective contact rates in community | Decrease | Tri(0.6, 1.0, 0.8) |
|  | [2020.25, 2022) | $\delta_{p} , \delta_{r} ,\delta_{d} , \delta_{n}$ | Diagnosis rates | Decrease | Calibrated |
| El Salvador | [2020.25, 2021) | $\beta_{cp}{,\beta}_{pc}, \beta_{cc}$ | Effective contact rates in community | Decrease | Tri(0.6, 1.0, 0.8) |
| Mexico | [2020.25, 2021) | $\beta_{cp}{,\beta}_{pc}, \beta_{cc}$ | Effective contact rates in community | Decrease | Tri(0.6, 1.0, 0.8) |
|  | [2020.25, 2022.5) | $\delta_{p} , \delta_{r} ,\delta_{d} , \delta_{n}$ | Diagnosis rates | Decrease | Calibrated |
| Peru | [2020.25, 2023) | $q_{r} , q_{d} , q_{n}$ | Prison entry rates | Decrease | Calibrated |
|  | [2020.25, 2021) | $\beta_{cp}{,\beta}_{pc}, \beta_{cc}$ | Effective contact rates in community | Decrease | Tri(0.6, 1.0, 0.8) |
|  | [2020.25, 2023) | $\delta_{p} , \delta_{r} ,\delta_{d} , \delta_{n}$ | Diagnosis rates | Decrease | Calibrated |

**Table S7. Relative values of calibrated within-prison tuberculosis parameters in 2019.** Ratios are shown for the values of each parameter in prison versus in the never incarcerated and distant incarceration history strata. Prison diagnosis rates are elevated in El Salvador due to a dramatic increase in notifications in prison starting in 2017. El Salvador’s national plan for tuberculosis control in 2017-2021 involved scaling up tuberculosis screening in prisons; therefore, we assume that the prison diagnosis rate spiked in those years, until the COVID-19 pandemic.

| Parameter | Argentina | Brazil | Colombia | El Salvador | Mexico | Peru |
| --- | --- | --- | --- | --- | --- | --- |
| Effective contact rate | 3.1 | 12.4 | 2.4 | 22.8 | 1.4 | 10.6 |
| Fast progression rate | 1.9 | 2.1 | 3.0 | 2.5 | 3.0 | 1.6 |
| Diagnosis rate | 0.4 | 0.6 | 0.5 | 1.3 | 0.6 | 0.4 |

**Table S8. Modeled percent change in prison entry and release rates from 1990-2019.** Percent change in entry rates represents the weighted percent change across all non-incarcerated strata, weighted by population proportion. Maximum changes are provided for countries that have partially reversed prior incarceration trends in recent years.

|  | Argentina | Brazil | Colombia | El Salvador | Mexico | Peru |
| --- | --- | --- | --- | --- | --- | --- |
| Net change in entry rates from 1990 to 2019 (%) | 514 (436, 592) | 501 (407, 574) | -15 (-40, 27) | 89 (72, 113) | -38 (-49, -26) | -18 (-26, -8) |
| Maximum change in entry rates between 1990 to 2019 (%) | Same | Same | 46 (13, 93) | 150 (125, 185) | 25 (5, 46) | 27 (20, 39) |
| Net change in release rates from 1990 to 2019 (%) | 40 (24, 55) | -4 (-18, 7) | -62 (-73, -45) | -63 (-70, -53) | -47 (-56, -37) | -78 (-81, -75) |
| Maximum change in release rates between 1990 to 2019 (%) | Same | -7 (-21, 0) | Same | Same | Same | Same |

**Table S9. Estimates of excess population tuberculosis incidence attributable to mass incarceration in 2022.** All estimates are at the population-level among individuals aged 15 and older. Incidence rate ratios and excess burden estimates were obtained from comparing incident tuberculosis cases between the observed scenario of mass incarceration and the counterfactual scenario of no change in incarceration prevalence since 1990. 95% uncertainty intervals are shown in parentheses.

|  | Incidence rate ratio for observed vs. counterfactual | Excess cases per 100,000 person-years relative to counterfactual | Absolute excess cases relative to counterfactual |
| --- | --- | --- | --- |
| Argentina | 1.09 (1.06, 1.21) | 2.4 (1.6, 5.4) | 832 (549, 1895) |
| Brazil | 1.51 (1.38, 1.7) | 15.9 (12.4, 21.3) | 27334 (21216, 36487) |
| Colombia | 1.24 (1.15, 1.4) | 6.2 (4, 9.9) | 2515 (1627, 4052) |
| El Salvador | 2.11 (1.85, 2.47) | 25.7 (19.8, 35.4) | 1215 (935, 1672) |
| Mexico | 1.06 (1.03, 1.09) | 1.4 (0.8, 2.4) | 1351 (749, 2352) |
| Peru | 1.23 (1.15, 1.33) | 22.7 (14.2, 31.9) | 5744 (3592, 8069) |

**Table S10. Results of multi-level meta-modeling.** Asterisks indicate confidence intervals that do not contain zero. Variables are listed in descending order by standardized coefficient estimate; variables with largest absolute magnitude of coefficient estimate are at the top and bottom of table. All variables representing parameters are baseline values unless indicated otherwise. SD, standard deviation; CI, confidence interval.

| Variable | Country-level SD | Standardized coefficient estimate (95% CI) |
| --- | --- | --- |
| Intercept | 0.025 | 1.4 (1.3, 1.4)* |
| Tuberculosis notification rate in prisons |  | 0.56 (0.38, 0.74)* |
| Release rate | 0.071 | 0.052 (-0.0059, 0.11) |
| Percent change in entry rates in **n** and **d** strata |  | 0.048 (0.013, 0.084)* |
| Incarceration prevalence in 2019 |  | 0.045 (-0.081, 0.17) |
| Diagnosis rate in **n** and **d** strata | 0.042 | 0.031 (-0.0041, 0.065) |
| Ratio of fast progression rate in **p** versus **n** and **d** strata |  | 0.022 (0.018, 0.027)* |
| Ratio of effective contact rate in **p** versus **n** and **d** strata |  | 0.019 (0.013, 0.024)* |
| TB mortality rate |  | 0.017 (0.015, 0.02)* |
| Percent change in diagnosis rate in **n** and **d** strata |  | 0.011 (0.009, 0.013)* |
| Weight for fast progression rate in **r** stratum |  | 0.0079 (0.0065, 0.0093)* |
| Entry rate in **n** and **d** strata |  | 0.0077 (-0.007, 0.022) |
| Relative risk of reinfection in L and R compartments |  | 0.0052 (0.0037, 0.0067)* |
| Rate of transition from early to late latent |  | 0.0036 (0.0017, 0.0056)* |
| Rate of self-cure |  | 0.0028 (0.0014, 0.0042)* |
| Mortality rate ratio in **r** stratum |  | 0.00054 (-0.00084, 0.0019) |
| Weight for diagnosis rate in in **r** stratum |  | 0.0005 (-0.00087, 0.0019) |
| Uncertainty factor for direct prison & community mixing |  | 0.00025 (-0.0011, 0.0016) |
| Mortality rate ratio in **p** |  | -0.00054 (-0.0019, 0.00083) |
| Ratio of entry rate in **r** versus **n** and **d** strata |  | -0.0092 (-0.014, -0.004)* |
| Percent change in release rate |  | -0.02 (-0.045, 0.005) |
| Fast progression rate in **n** and **d** strata | 0.025 | -0.034 (-0.056, -0.012)* |
| Percent change in effective contact rate in **r**, **n**, and **d** strata | 0.093 | -0.082 (-0.16, -0.0065)* |
| Effective contact rate in **r**, **n**, and **d** strata | 0.068 | -0.092 (-0.15, -0.037)* |
| Population tuberculosis notification rate |  | -0.33 (-0.41, -0.24)* |

**Table S11. Percent of excess incident cases in 2019 among formerly incarcerated individuals.** 95% uncertainty intervals are shown in parentheses.

|  | Recent incarceration history (%) | Distant incarceration history (%) | Any incarceration history (%) |
| --- | --- | --- | --- |
| Argentina | 23 (16, 30) | 11 (7, 16) | 34 (24, 45) |
| Brazil | 27 (19, 34) | 8 (6, 10) | 34 (26, 42) |
| Colombia | 13 (7, 18) | 3 (2, 5) | 16 (10, 22) |
| El Salvador | 10 (6, 15) | 2 (1, 2) | 11 (6.8, 17) |
| Mexico | 15 (9, 22) | 11 (6, 16) | 26 (16, 36) |
| Peru | 4 (0, 7) | 0 (0, 1) | 4 (0, 8) |

**Table S12. Model outputs for stratum-specific tuberculosis incidence rates per 100,000 person-years in 2019.** Median estimates are shown with 95% uncertainty intervals in parentheses.

|  | Total (population-wide) | Prison | Recent incarceration history | Distant incarceration history | Never incarcerated |
| --- | --- | --- | --- | --- | --- |
| Argentina | 27 (24-30) | 487 (367-1148) | 140 (96-291) | 86 (77-112) | 24 (22-27) |
| Brazil | 46 (43-52) | 2094 (1666-2790) | 559 (355-837) | 131 (112-153) | 30 (26-34) |
| Colombia | 32 (28-38) | 1503 (949-2487) | 213 (116-386) | 61 (53-81) | 25 (21-30) |
| El Salvador | 57 (48-67) | 3198 (2474-4132) | 656 (410-952) | 90 (77-106) | 24 (21-27) |
| Mexico | 23 (19-29) | 369 (245-564) | 66 (43-102) | 42 (37-49) | 21 (17-26) |
| Peru | 118 (98-141) | 5445 (3582-9025) | 892 (616-1496) | 168 (156-180) | 91 (75-109) |

**Table S13. Incarceration prevalence in 2034 under future incarceration scenarios.** Prevalence is shown per 100,000 among those aged 15 and older. 95% uncertainty intervals are provided in parentheses. All changes in entry rates and duration occur gradually between 2024-2034, reaching the percentage target by 2034. Future projections for El Salvador were performed separately.

|  | Stable entry & release rates | Continue trends | Reduce entry rates by 25% | Reduce entry rates by 50% | Reduce duration by 25% | Reduce duration by 50% | Reduce entry rates & duration by 25% | Redyce entry rates & duration by 50% |
| --- | --- | --- | --- | --- | --- | --- | --- | --- |
| Argentina | 392 (371-411) | 469 (451-490) | 315 (295-336) | 240 (221-262) | 322 (300-344) | 231 (211-254) | 255 (235-278) | 128 (114-147) |
| Brazil | 473 (458-484) | 461 (421-493) | 361 (351-369) | 253 (245-263) | 377 (368-386) | 265 (257-274) | 283 (275-292) | 130 (124-137) |
| Colombia | 307 (295-317) | 270 (252-290) | 251 (242-257) | 196 (188-202) | 252 (242-257) | 179 (172-183) | 203 (195-208) | 104 (99-111) |
| Mexico | 242 (231-260) | 217 (197-244) | 191 (182-207) | 142 (133-155) | 191 (182-206) | 131 (125-142) | 149 (141-162) | 71 (66-78) |
| Peru | 418 (392-435) | 458 (423-479) | 352 (329-368) | 286 (266-305) | 357 (337-373) | 270 (254-287) | 297 (279-315) | 170 (155-190) |

**Table S14. Additional parameters to model El Salvador’s state of emergency (SoE).**

| Parameter | Description | Calibrated or sampled? | Prior [box constraints if calibrated] | | Source / rationale |
| --- | --- | --- | --- | --- | --- |
| SoEf | Multiplier on pre-SoE $q_{d}$ and$q_{n}$for March-June 2022 | Calibrated | | [20, 60] | Based on rate of arrests in the first three months^18^, compared to pre-SoE |
| SoEf_iR | Multiplier on pre-SoE $q_{r}$ relative to *SoEf*, for March-June 2022 | Sampled | | Unif(0.25, 1.25) | No data to inform assumption. Translates to people w/ incarceration history comprising ~10-25% of arrested individuals during SoE |
| SoE_r | Release rate from March 2022-January 2024 | Sampled | | Unif(0.065, 0.1) | Based on statements about number released among new detainees^20,21^, with upper bound representing additional, as-usual releases of those detained pre-SoE |
| SoEf2 | Multiplier on pre-SoE $q_{r}, q_{d}$, $q_{n}$ for August 2023-present | Calibrated | | [1.5, 4] | Based on rate of arrests since August 2023^18^, compared to pre-SoE |

**Table S15. Incarceration prevalence in 2034 under future incarceration scenarios in El Salvador.** Prevalence is shown per 100,000 among those aged 15 and older. 95% uncertainty intervals are provided in parentheses.

|  | Continue state of emergency | Gradual passive abatement | Active 10-year reversion | Active 5-year reversion | Active 2-year reversion |
| --- | --- | --- | --- | --- | --- |
| El Salvador | 2777 (2162-3317) | 1604 (1382-1810) | 842 (707-1036) | 508 (411-674) | 299 (245-371) |

**Figure S1. Schematic of meta-population dynamic compartmental model.** S, susceptible; E, early latent infection; L, late latent infection; I, infectious active disease; R, recovered. Lowercase subscripts represent population strata: p, prison; r, recent history of incarceration; d, distant history of incarceration; n, never incarcerated. Black solid lines represent natural history transitions. Purple dashed lines represent transitions across population strata. Green and red lines represent births and deaths, respectively. Parameters are defined in **Table S3**. For reinfections from R🡪E or L🡪E, $\beta I$ represents a simplified annotation of the force of infection for a Susceptible individual.

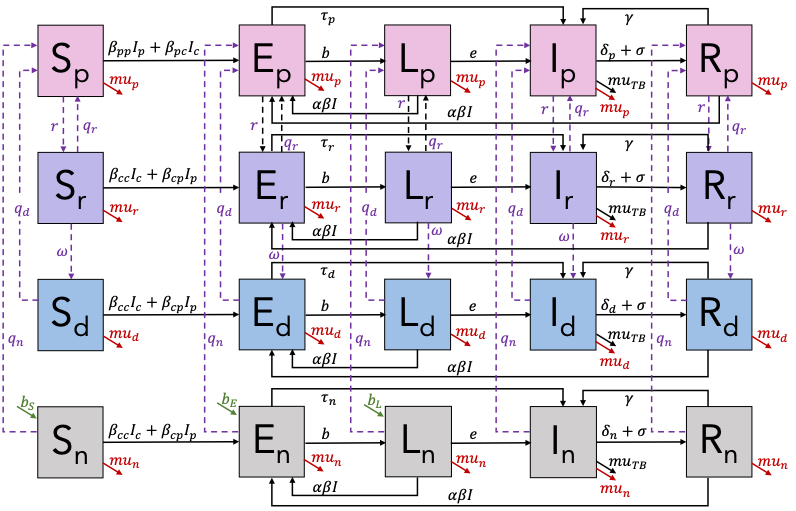

**Figure S2. Posterior distribution for regional case detection ratio (CDR) in prisons.**

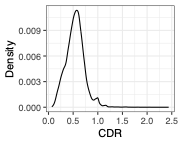

**Figure S3. Schematic of simple population transition model for calibrating incarceration-related parameters.** Population strata include: p1, first time incarcerated; p2, repeat incarcerated; r, recent history of incarceration; d, distant history of incarceration, n, never incarcerated. Parameters include: *q_i_*, prison entry rate from stratum *i*; r, release rate; $\omega$, rate of transition from recent to distant incarceration history. Individuals are born into the never incarcerated stratum (birth and death rates not shown).

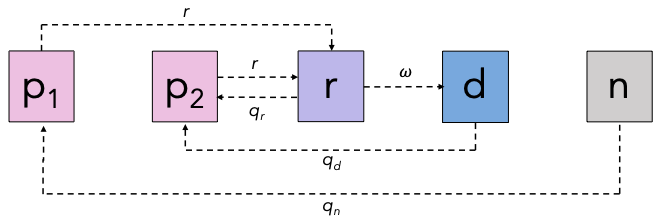

**Figure S4. Prior and posterior distributions for calibrated model parameters.** Prior and posterior distributions are shown in blue and red, respectively. For time-varying parameters, values at baseline are shown, with changes over time shown in **Figure S5.** Text shows means, with 95% intervals in parentheses. q_i, prison admissions rate in stratum i; r, prison release rate; beta_pp, effective contact rate for transmission within prison; beta_cc, effective contact rate for transmission outside prison; tau_i, fast progression rate in stratum i, delta_i, diagnosis rate in stratum i.

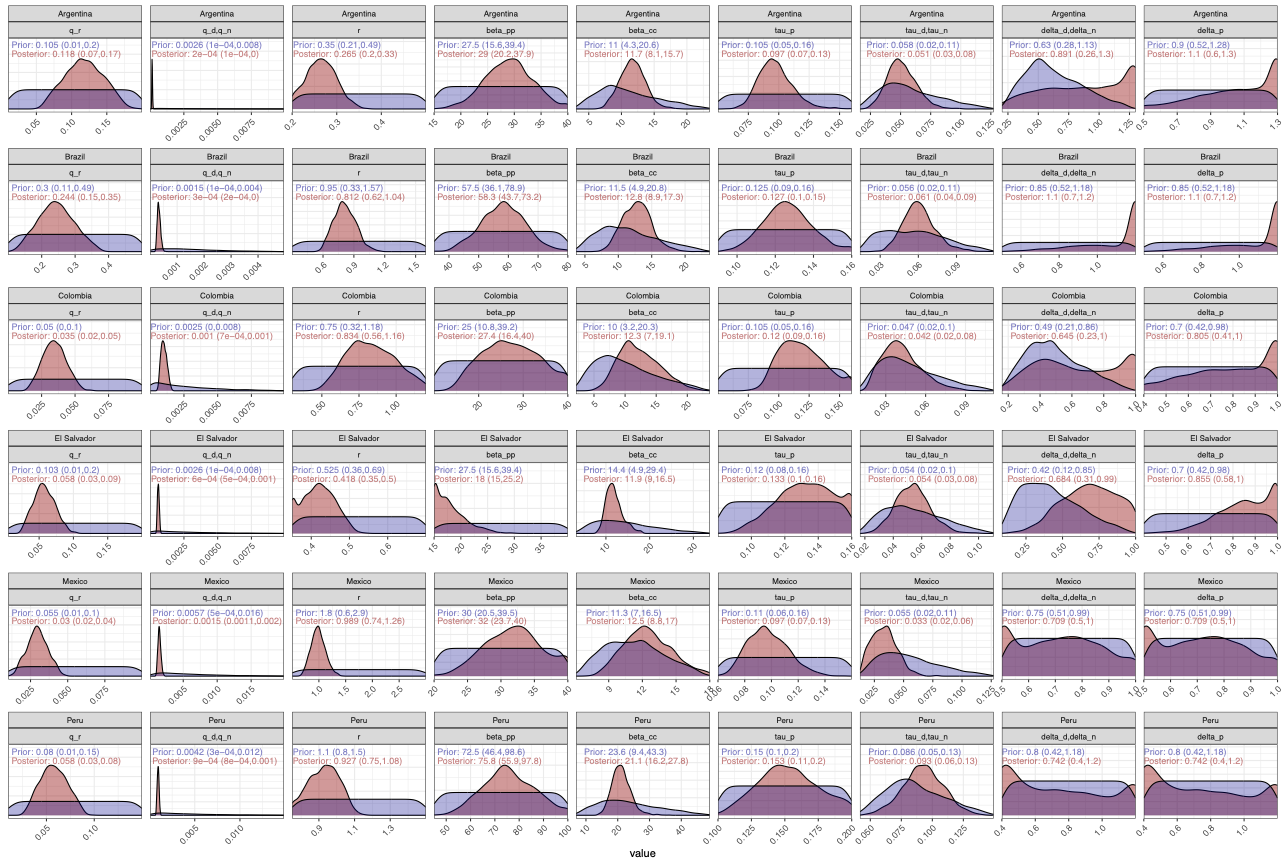

**Figure S5. Time-varying parameters.** Posterior medians and 95% intervals are shown in black lines and grey shaded bands, respectively. q_i, prison admissions rate in stratum i; r, prison release rate; beta_cc, effective contact rate for transmission outside prison; beta_pp, effective contact rate for transmission within prison; delta_i, diagnosis rate in stratum i; mu_i, mortality rate in stratum i; vi, ratio of birth rate to death rate.

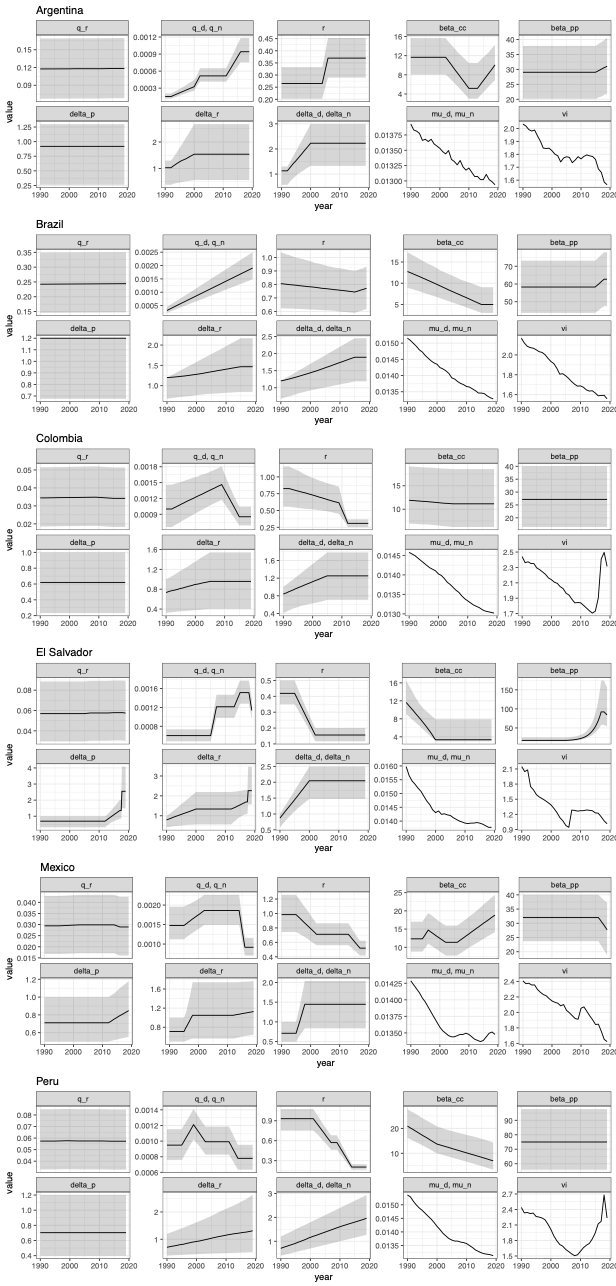

**Figure S6. Model fit to incarceration-related calibration targets.** Black points and error bars represent calibration targets and 95% uncertainty bounds (if applicable), respectively. Dark blue lines and shaded bands represent median model fits and 95% uncertainty intervals, respectively. In Brazil, recidivism data was only available for one year (2013); the calibration target and uncertainty bounds are shown by the vertical black and dotted lines, respectively. Incarc prev, incarceration prevalence; 100k, 100,000 population age 15+.

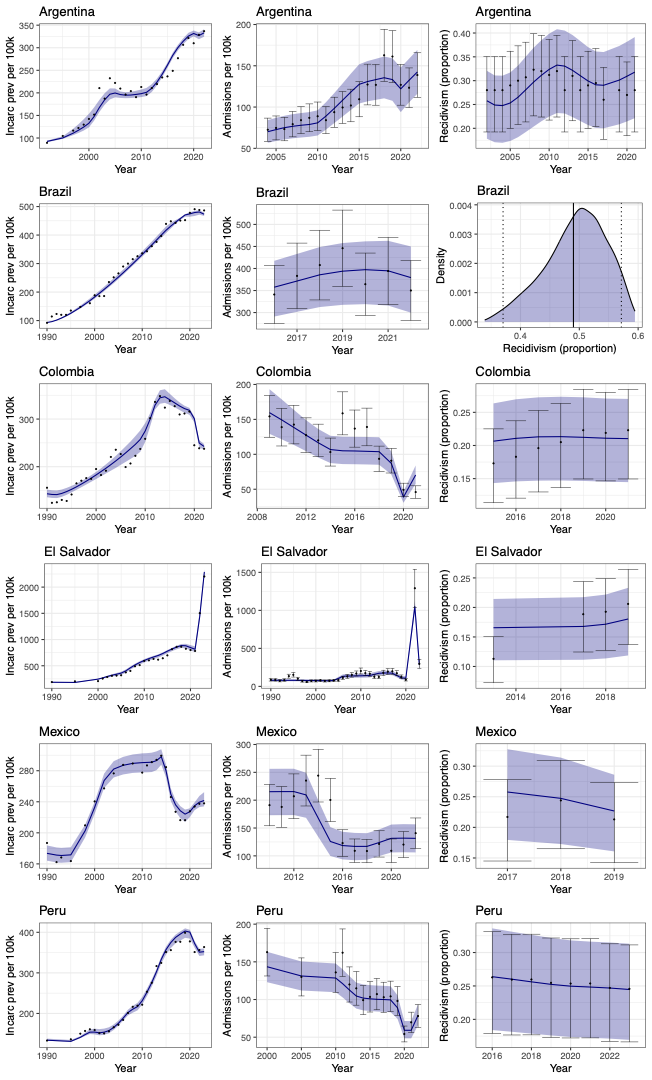

**Figure S7. Model fit to tuberculosis-related calibration targets.** Black points and error bars represent calibration targets and 95% uncertainty bounds (if applicable), respectively. Dark blue lines and shaded bands represent median model fits and 95% uncertainty intervals, respectively. “Combined” indicates population-level incidence and notifications. 100k, 100,000 person-years.

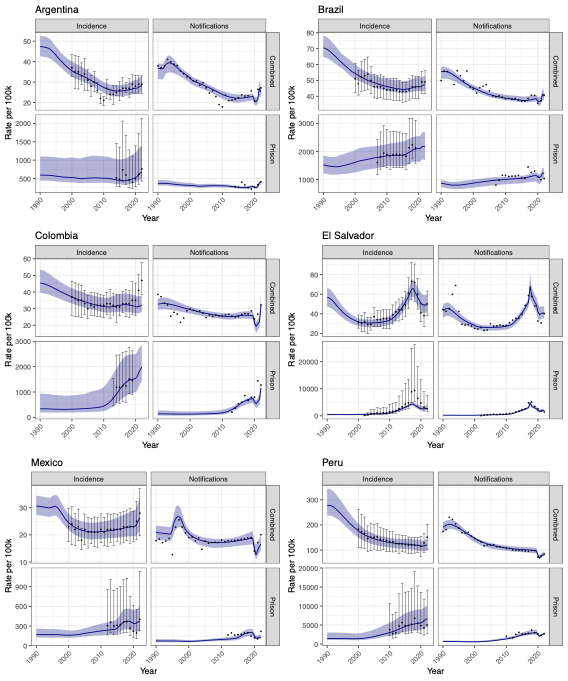

**Figure S8. Model consistency with validation data.** Data used for validation are shown in black points and error bars. Dark blue lines and shaded bands represent median model outputs and uncertainty intervals for A) the population-level rate of tuberculosis deaths per 100,000 person-years, B) the prevalence of tuberculosis in prison, and C) the prevalence of latent tuberculosis infection (LTBI) in prison. Data on B) and C) were only available for Brazil and Colombia.

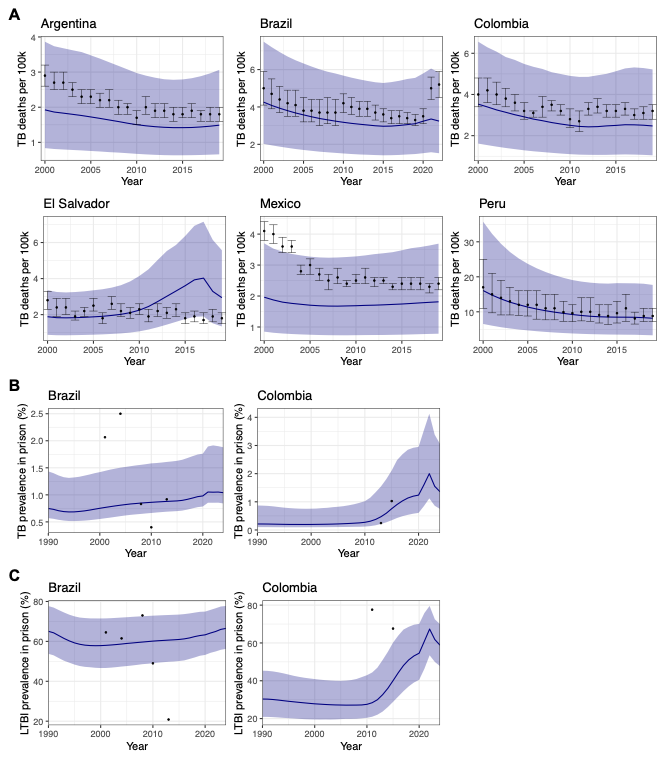

**Figure S9. Results from five sensitivity analyses.** Incidence rate ratios were obtained from comparing incident tuberculosis cases between the observed scenario of the historical rise in incarceration and the counterfactual scenario of no change in incarceration prevalence since 1990. Parameters were re-calibrated when necessary to achieve a sufficient fit to the data, given different assumptions in each sensitivity analysis. Sensitivity analyses were as follows: 1) 1:2 ratio of early to late latent infection among individuals born into the model at age 15 with latent infection; 2) equivalent fast progression rate (tau) and diagnosis rate (delta) in recent incarceration history stratum as in distant history and never incarcerated strata; 3) no changes over time in the effective contact rate for within-prison mixing, beta_pp; 4) assortative mixing among non-incarcerated strata based on incarceration history; 5) no mixing between incarcerated and non-incarcerated individuals. Sensitivity analysis #3 was not performed for Colombia and Peru, where beta_pp did not change over time in the main analysis. In El Salvador, we were unable to achieve an adequate fit to calibration targets without changing beta_pp over time.

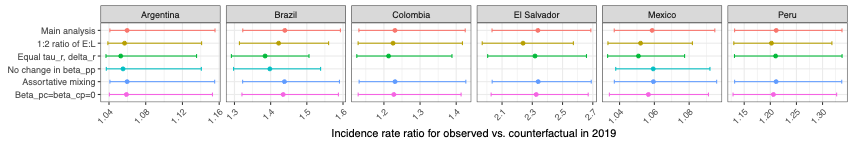

**Figure S10. Stratum-specific tuberculosis incidence rates.** Model outputs for tuberculosis (TB) incidence rates per 100,000 person-years in the total population and in each stratum in the year 2019. The y-axis is on a log10 scale. Outliers are not shown. Incarc., incarceration.

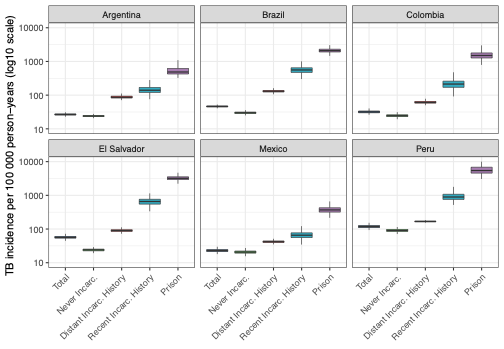

**Figure S11. Projected impacts of El Salvador’s state of emergency on population TB incidence.**
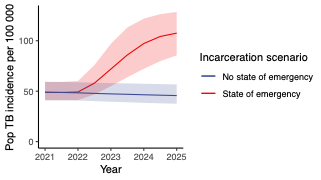

20.	El Salvador libera a 7.000 personas que habían sido detenidas durante el régimen de excepción. Europa Press Internacional. 2023 August 23, 2023.

21.	El Salvador libera a 3.000 detenidos en la guerra contra pandillas, dice Bukele. France 24. 2023 January 18, 2023.

22.	Sistema Nacional de Estadísticas sobre Ejecución de la Pena (SNEEP). Informes SNEEP. <https://www.argentina.gob.ar/justicia/politicacriminal/estadisticas/sneep>.

23.	Sistema de Informações do Departamento Penitenciário Nacional (SISDEPEN). Dados Estatísticos do Sistema Penitenciário. <https://www.gov.br/senappen/pt-br/servicos/sisdepen>.

24.	Sanguinetti P, Ortega D, Berniell L, et al. RED 2014: Por una América Latina más segura: Una nueva perspectiva para prevenir y controlar el delito: Banco de Desarrollo de América Latina (CAF), 2014.

25.	Bergman M, Amaya LE, Fondevila G, Vilalta C. Reporte de cárceles en El Salvador: Perfiles generales, contexto familiar, delitos, circunstancias del proceso penal y condiciones de vida en la cárcel: Universidad Francisco Gavida (UFG), 2015.

26.	Claudia Cifali A, de Azevedo RG. Public Security, Criminal Policy and Sentencing in Brazil during the Lula and Dilma Governments, 2003-2014: Changes and Continuities. *International Journal for Crime, Justice and Social Democracy* 2017; **6**(1): 146-63.

27.	Instituto Nacional Penitenciario y Carcelario (INPEC). Informes y Boletines Estadísticos. <https://www.inpec.gov.co/en/estadisticas/informes-y-boletines>.

28.	Dirección General de Centros Penales (DGCP).

29.	Andrade L, Carrillo A. El sistema penitenciario salvadoreño y sus prisiones: Instituto Universitario de Opinión Pública (IUDOP); Universidad Centroamericana “José Simeón Cañas”, 2015.

30.	Arévalo M. Aumentan reincidencias en sistema penitenciario. *La Prensa Gráfica*, August 7, 2019, 2019. <https://www.laprensagrafica.com/elsalvador/Aumentan-reincidencias-en-sistema-penitenciario-20190806-0433.html> (accessed.

31.	Instituto Nacional de Estadística y Geografia (INEGI). <https://en.www.inegi.org.mx/programas/cngspspe/>.

32.	Instituto Nacional Penitenciario (INPE). Sistema de Información Estadístico Penitenciario (SIEP). <https://www.inpe.gob.pe/estad%C3%ADstica1.html>.

33.	Magán Zevallos JC. Overcrowding in the Peruvian prison system. International Review of the Red Cross: International Committee of the Red Cross, 2016.

34.	Menzies NA, Wolf E, Connors D, et al. Progression from latent infection to active disease in dynamic tuberculosis transmission models: a systematic review of the validity of modelling assumptions. *Lancet Infect Dis* 2018; **18**(8): e228-e38.

35.	Andrews JR, Noubary F, Walensky RP, Cerda R, Losina E, Horsburgh CR. Risk of progression to active tuberculosis following reinfection with Mycobacterium tuberculosis. *Clin Infect Dis* 2012; **54**(6): 784-91.

36.	Blower SM, McLean AR, Porco TC, et al. The intrinsic transmission dynamics of tuberculosis epidemics. *Nature Medicine* 1995; **1**(8): 815-21.

37.	Ragonnet R, Flegg JA, Brilleman SL, et al. Revisiting the Natural History of Pulmonary Tuberculosis: A Bayesian Estimation of Natural Recovery and Mortality Rates. *Clin Infect Dis* 2021; **73**(1): e88-e96.

38.	World Population Prospects 2022. In: Nations U, editor. 27 ed; 2022.
